# The interpupillary distance differs between ethnicities and associates with horizontal strabismus patterns: Evidence from a systematic review and meta-analysis

**DOI:** 10.64898/2025.12.30.25343217

**Authors:** Zainab Zehra, Molly M. Hagen, Lingchen Wang, Christopher S. von Bartheld

## Abstract

**Purpose:** The interpupillary distance is a measure of the width of the orbit and is important for spectacle design and proper head-mounted displays in virtual reality. Extreme interpupillary distances may predispose to horizontal strabismus. The interpupillary distance is thought to differ between ethnicities, but global data on this parameter have not been systematically explored, mapped and compared.

**Methodology:** We performed a systematic review that compiled 242 studies reporting the interpupillary distance and mapped the data geographically. We then compared the regional distribution of the mean interpupillary distance with the distribution of horizontal strabismus patterns. The strabismus data, obtained from our previous systematic review of the literature, were used to map the relative frequency of esotropia and exotropia according to 301 population-based studies. The mean interpupillary distance and esotropia/exotropia ratio of major ethnicities were then analyzed by meta-regression analyses to determine an association between the two parameters.

**Results:** Interpupillary distances are larger in Western Africa, South India, East Asia, in Latinos/Hispanics, in Native and African Americans, and they are smaller in Europe, North Africa, the Middle East, Northwestern India, and in Inuit populations. Regression analyses revealed an association between the interpupillary distance and the esotropia/exotropia ratio with R^2^ values of 0.320 (major ethnicities) and 0.410 (populations at higher resolution).

**Conclusion:** The mean interpupillary distance varies significantly between ethnicities. Orbital anatomical parameters contribute to diverse horizontal strabismus patterns. Our findings may aid in the design of appropriate spectacles and the optimal size of head-mounted displays, and help to better understand the pathogenesis of horizontal strabismus.

Graphical Abstract

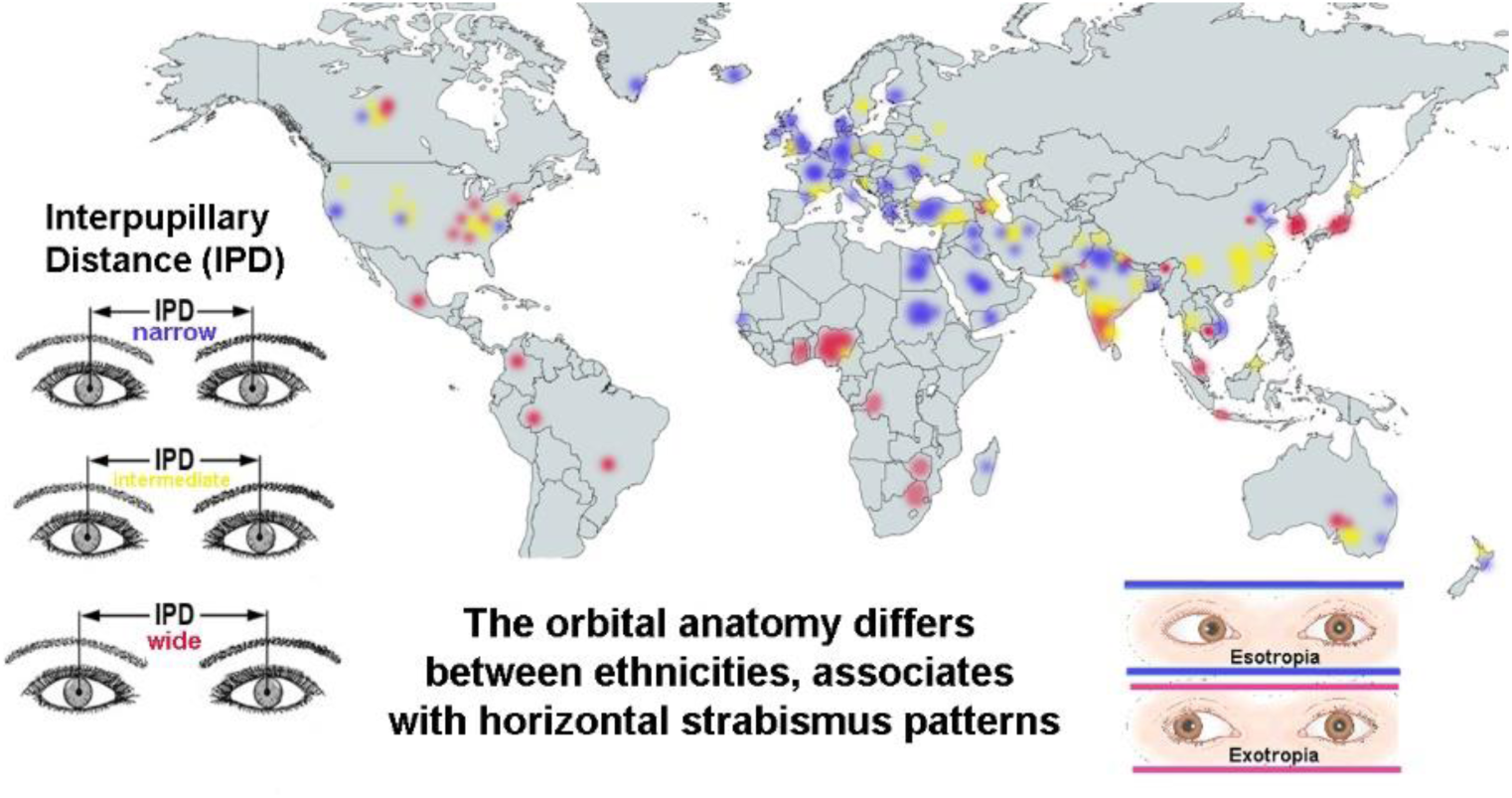

## Introduction

The interpupillary distance (IPD) is the distance between the centers of the right and left pupil and can be used as an approximate measure of the orbital width.^1–2^ The mean IPD differs between ethnicities, a notion that is based on a relatively small number of studies from select populations.^3–4^ Despite many studies reporting the mean IPD in cohorts, global data on this parameter have not been systematically explored, mapped and compared. Numerous studies from dental schools examined and reported the IPD because of the importance of this craniofacial parameter for proportionality of maxillary anterior teeth.^5–8^

The IPD is important for spectacle design^9–11^ and for suitable head-mounted displays in virtual reality.^12–17^ A mismatch between the user’s IPD and the IPD on the head-mounted display can cause discomfort.^18^ Inadequate ranges of such headsets are thought to be responsible for the female propensity to cybersickness.^16^ In fact, cybersickness seen in virtual reality is thought to be due, largely, to an improper orbital range of the headsets.^16,18^

Differences in the IPD have been proposed to explain ethnic differences in horizontal strabismus patterns.^19–20^ Recent studies provided evidence for surprisingly distinct and consistent ethnic differences in the prevalence and the patterns of horizontal strabismus, with some populations having much more esotropia than exotropia, while other populations have more exotropia than esotropia.^21–22^ Historically, the orbital anatomy, and especially the IPD, was suspected to play a role in strabismus, as first suggested in the 19^th^ century.^19–20,23–31^ However, this line of research has not been pursued for over 80 years.^22^ A large number of population-based studies has now been conducted and published, both on the global distribution of the IPD,^32^ and on the global distribution of esotropia and exotropia.^22^ Such data make it possible to map these parameters across populations and to determine an association between them at the population level. Does the orbital width explain, in part, why horizontal strabismus patterns vary between different populations?

Our systematic review and meta-analysis compiles information about the IPD as a proxy of the orbital width. We mapped the mean IPD side-by-side with strabismus patterns in different populations based on studies from 61 countries throughout the world. This may aid the design of headset ranges suitable for diverse populations. We show that a larger IPD associates with a lower esotropia/exotropia ratio, as previously hypothesized.^19–20,30,33–34^ Our work is the first to provide evidence for an association between the IPD and horizontal strabismus patterns at the population level. These new insights contribute to a better understanding of the mechanisms underlying diverse geographical patterns of horizontal strabismus.

## Material and methods

To compile data on the orbital width, we conducted a systematic review of studies by searching Google Scholar with the key words “interpupillary distance” and “IPD,” last updated on 29 November, 2024 (Fig. 1). The references within relevant studies were examined for additional eligible sources. We included only data from adults and from teenagers when they had reached adult values of the IPD or interorbital width (Supplemental Table 1).^35–43^ Studies that explicitly acknowledged biased data collection were excluded.^44^

**Figure 1:**
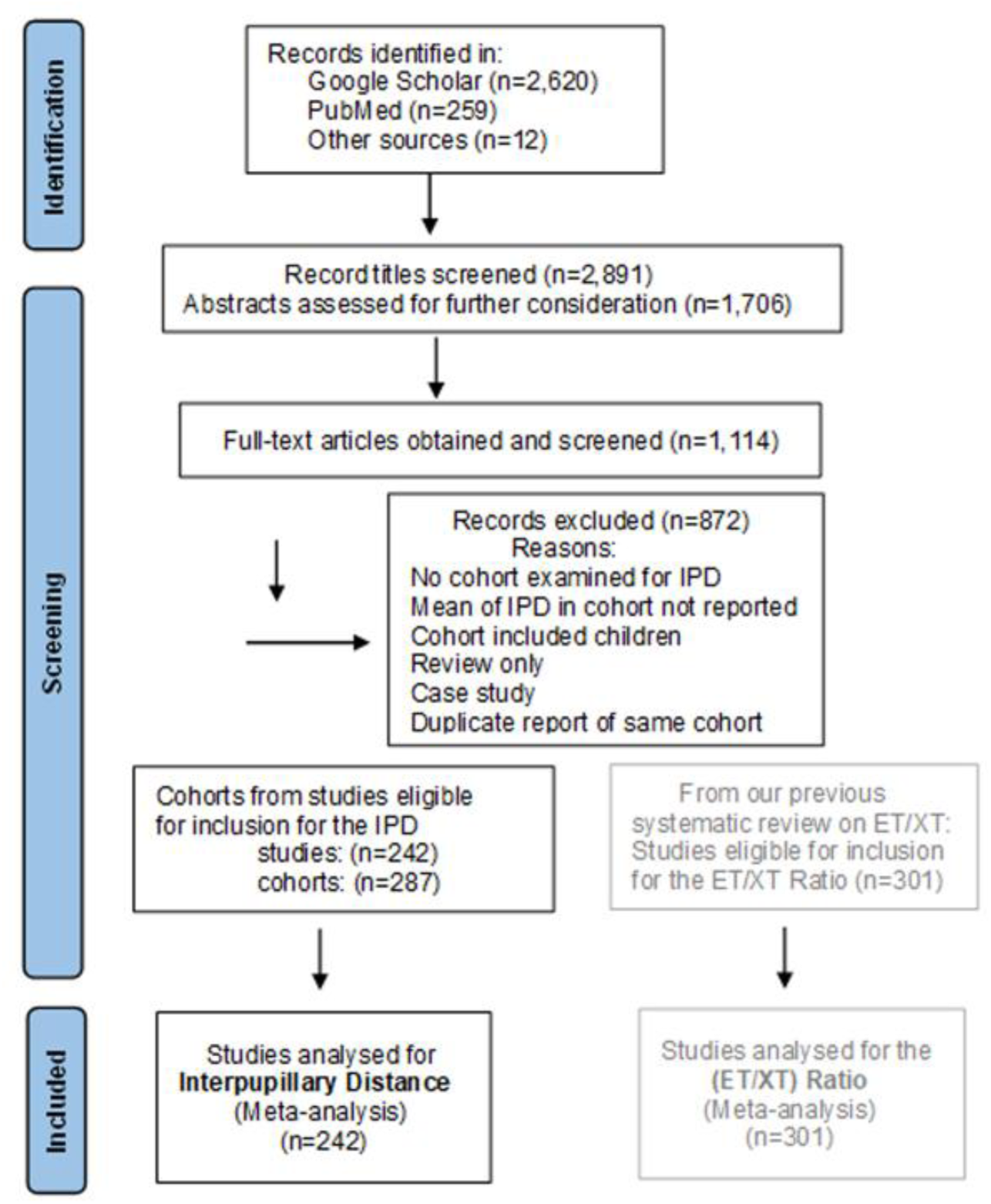
PRISMA Flowchart of the search and screening strategy for studies on the interpupillary distance (IPD). The searches for the literature on the IPD and the horizontal strabismus were last updated on November 29, 2024. The studies eligible for inclusion for the esotropia/exotropia ratio (ET/XT ratio) were obtained from our previous systematic review and meta-analysis^22^ (for the 301 references on the ET/XT ratio, see Supplemental Table S3).

The studies reporting strabismus prevalence were taken from our previous systematic reviews and meta-analyses.^22,45^ We only included studies that reported numerical values on prevalence of esotropia and exotropia. We excluded studies that were limited to subjects 2 years of age or younger, because strabismus often develops after 2 years of age.^46–51^ We excluded clinic-based studies, case reports, and we removed duplicate studies reporting on the same cohorts.

We created two types of geographical maps, the first one shows the global distribution of the mean IPD, and the second one shows the distribution of horizontal strabismus patterns, based on representative samples of the populations. Maps of the mean IPD were designed by using a graded heat map with color-coding of narrow (<61 mm, blue), intermediate-narrow (61-62 mm, green), intermediate (62-63 mm, yellow), relatively wide (63-64 mm, brown), and very wide distances (>64 mm, red). The heat maps were adjusted in the ranges to optimize visualization of differences and gradients between populations. National data (subjects in cohorts comprised from the entire country) are indicated in the center of the country. Ethnicities other than the predominant one of the country are indicated with special symbols. Likewise, cohorts of people with Down syndrome are coded separately.

Strabismus maps were created by calculating the esotropia/exotropia ratio (ET/XT ratio) for each study or cohort. We plotted the ET/XT ratio in a graded heat map with a lower than 0.25 ratio in red, 0.25 to 0.75 ratio in brown, 0.75 to 1.25 ratio in yellow, 1.25 to 3.0 ratio in green, and higher than 3.0 in blue. Ratios were adjusted in the ranges for optimal visualization of gradients. The size of the symbols is representative of the size of the cohort, with different symbols (circle, square, triangle, star, pentagon, rhombus) indicating different ethnicities (in countries with multiple ethnicities when ethnicity-specific information was reported). The ET/XT ratios for small cohorts were adjusted to avoid having a “0” in the numerator or in the denominator (Supplemental Table S2).^52^

We calculated mean IPDs and ET/XT ratios for the different regions and major ethnicities by adding up the mean times the sample size for each sample, and dividing this number by the sum of the sample sizes. We used these data to conduct a bivariate regression analysis that measured the strength of correlation between the IPD and the ET/XT ratio. Since some studies examined only male IPDs, some studies examined only females IPDs, and some studies reported only the mean IPD of males and females combined, we estimated any missing values by using a consistent formula for each ethnicity, as indicated in the Supplemental Table S1. The mean of the male value plus the mean of the female value divided by 2 was used for the overall IPD estimate. The IPD data were plotted against the ET/XT ratio of the same population, region or ethnicity. We pursued two strategies: pairing the IPD and ET/XT values only for major ethnicities, as well as pairing values derived at a higher resolution, from cohorts within 81 regions where data was available for both parameters (Supplemental Table S4). Outliers were identified using Tukey’s Hinges with k=3.0 and were excluded from the regression analysis.

We also used random effects meta-analysis to calculate a pooled mean IPD estimate for each ethnicity that incorporated between-study variation. We used mean and standard deviation data from each cohort with the meta-gen function with the inverse variance method for pooling from the R-meta package, version 4.9-5, in R software (R Foundation for Statistical Computing, Vienna, Austria) to estimate mean pooled IPD with 95% confidence intervals.^53^ Approximately 77% of the cohorts (n=222/287) in this review included standard deviation data (or data from which it could be calculated), and were eligible to be included in the meta-analyses. Heterogeneity across studies was assessed using the Maximum-likelihood estimator, Higgins’ I2 and Cochran’s Q method. We used a subgroup test to examine differences in pooled random IPD between participants of different ethnicities and estimate 95% confidence intervals for each group. Ethnic groups with at least three studies were included in the meta-analysis. Significance was defined at p=0.05.

## Results

### 1. Mapping of the interpupillary distance (IPD)

Eligible studies were mapped geographically by the size of the cohort. Only data from adults or teenagers 14 years and older were included, except for Down syndrome cohorts where adult IPDs are reached at an earlier age.^54–57^ We report on the mean IPD from n=242 studies reporting on 125,973 subjects in 287 cohorts.^32^

a. Europe. We found 42 studies reporting on 48 cohorts, with a total number of 15,968 subjects in all cohorts combined.^1,9,12–14,17,58–88^ The geographical map for Europe is shown in Fig. 2A. Most of the mean IPDs in Central Europe range from ∼ 60 mm to 63 mm, with larger mean IPDs (62-65 mm) present in Eastern Europe, Eastern Turkey and the South Caucasus.
b. Africa. We found 27 studies reporting on 31 cohorts with 13,169 subjects.^30,42,69,82,89–111^ Two basic patterns are present: North and East Africa has populations with a narrower IPD of 61-64 mm, while West Africa has populations with the widest IPD, of 65 to over 70 mm (Fig. 3A).
c. Middle East. We found 36 studies reporting on 35 cohorts with 15,041 subjects.^10,112–145^ Most studies from this region report a narrow (<63 mm) IPD, including Arabs, Jews, some Iranians, and most Turks. In Eastern Turkey and Azerbaijan, the mean IPD is higher with 63 to 67 mm (Fig. 3C).
d. Asia and Oceania. We found 50 studies reporting on 51 cohorts with 13,980 subjects in South Asia,^11,43,57,82,146–191^ 46 studies with 25,041 subjects in 56 cohorts from East Asia,^18,36,38,40,82,184,192–231^ and 7 studies with 1,925 subjects in 11 cohorts from Oceania, including Australia^232–238^ (Fig. 4A). The studies from the Northwest of the Indian subcontinent mostly showed a narrow IPD of less than 61 mm, while Central India and especially the South of India had larger IPDs of 61 to over 65 mm. The studies from East Asia report IPDs of 61 to 63 mm in China, and larger IPDs in Japan and Korea, mostly 63 to over 65 mm. For Oceania, most studies reported on Caucasians or indigenous populations (Maoris and Samoans).^232^
e. Americas. We found 52 studies reporting on 54 cohorts with 40,773 subjects; 46 studies were from North America (38,884 subjects),^4–7,29,35–36,41,82,239–263^ and 6 studies from South and Central America (1,889 subjects).^36,82,264–268^ The studies from North and Central America included 28 studies with 26,576 Caucasian subjects, 11 studies with 3,902 African Americans, 3 studies from North America on multiethnic cohorts (7,503 subjects), 9 studies from North and South America with 2,766 Latinos/Hispanics, and one study on 26 Native Americans. As expected for a continent with multiple ethnicities, the five major ethnicities (Caucasian, Latino/Hispanic, African American, Native American, Inuit/Eskimo) show a large diversity of mean IPDs. Inuit (Eskimos) have a very narrow IPD of 59-60 mm,^88^ while Native Americans have a wide IPD of over 65 mm.^4^ Most studies on Caucasians in North America report an IPD similar to Caucasians in Europe. African Americans have a wider IPD, of 64-68 mm, which is similar to Africans from West Africa. In South and Central America, most studies on Latinos/Hispanics report an intermediate to wide IPD of 63-64 mm.

**Figure 2:**
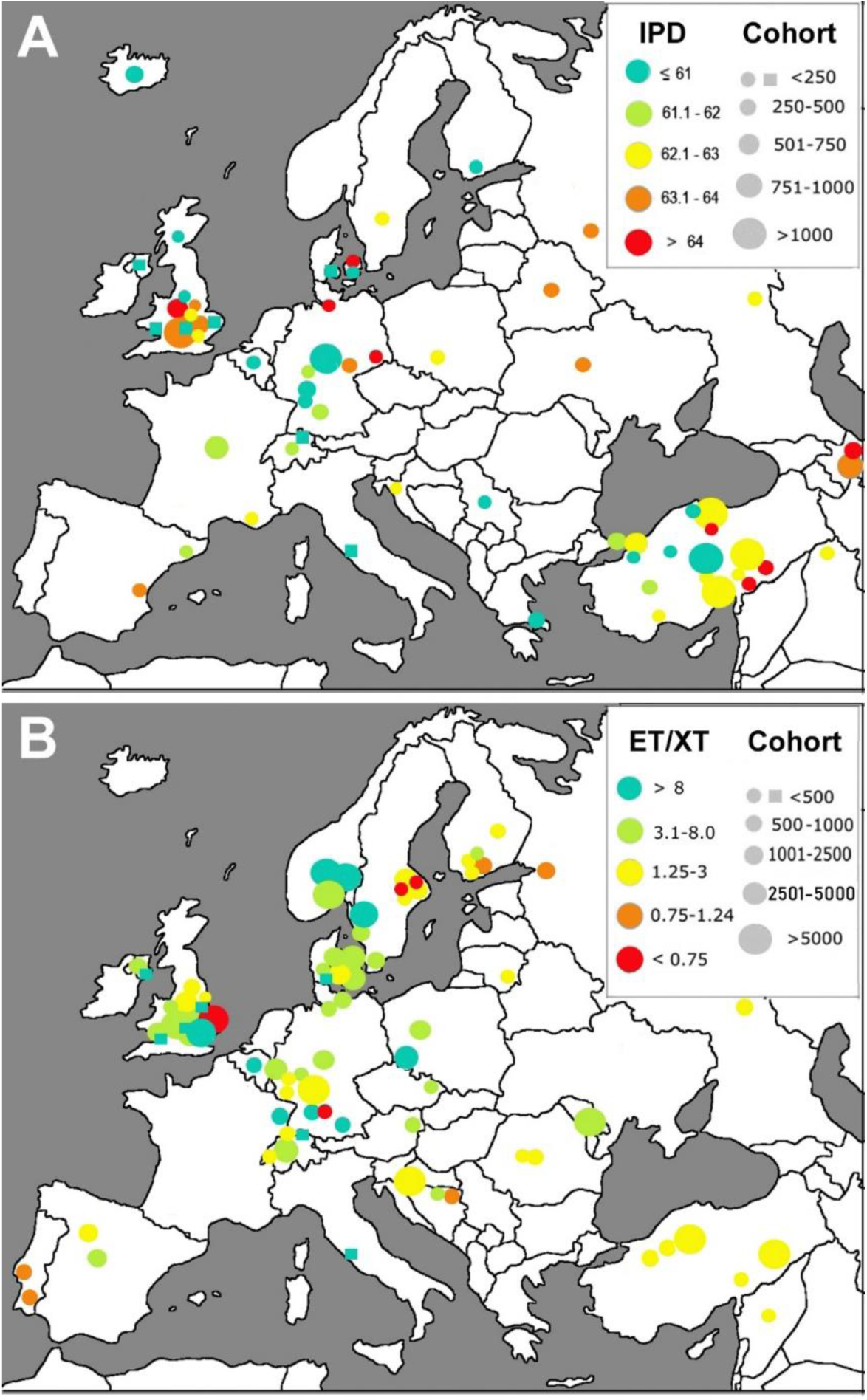
Europe. Geographical mapping of the interpupillary distance (IPD) (A) and the esotropia/exotropia ratio (ET/XT ratio) (B) in Europe. The size of the circles denotes the size of the cohort examined. The ranges of the IPD and the ET/XT ratio are color-coded in a heat map as indicated. Cohorts of people with Down Syndrome are shown as squares. Data from one cohort of Inuit (Eskimos) in Greenland is shown in Fig. 5A. National data (subjects in cohorts comprised from the entire country) are indicated in the center of the country.

**Figure 3:**
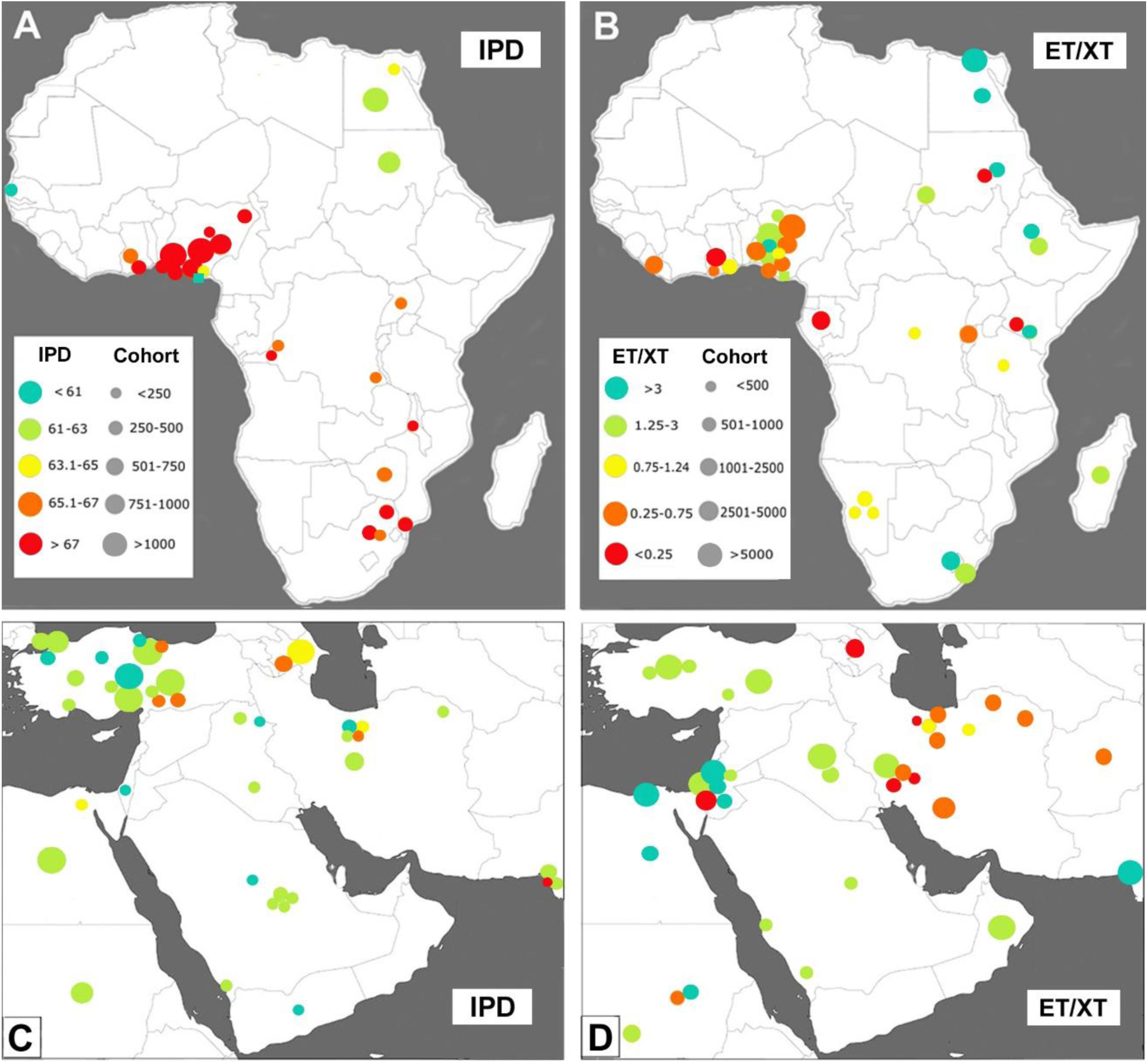
Africa and Middle East. Geographical mapping of the interpupillary distance (IPD) (A) and the esotropia/exotropia ratio (ET/XT ratio) (B) in Africa, and mapping of the IPD and ET/XT ratio (C, D respectively) in the Middle East. The size of the circles denotes the size of the cohort examined, and the ranges of the IPDs and the ET/XT ratios are color-coded in a heat map as indicated in panels A for the IPD, and panel B for the ET/XT ratio.

**Figure 4:**
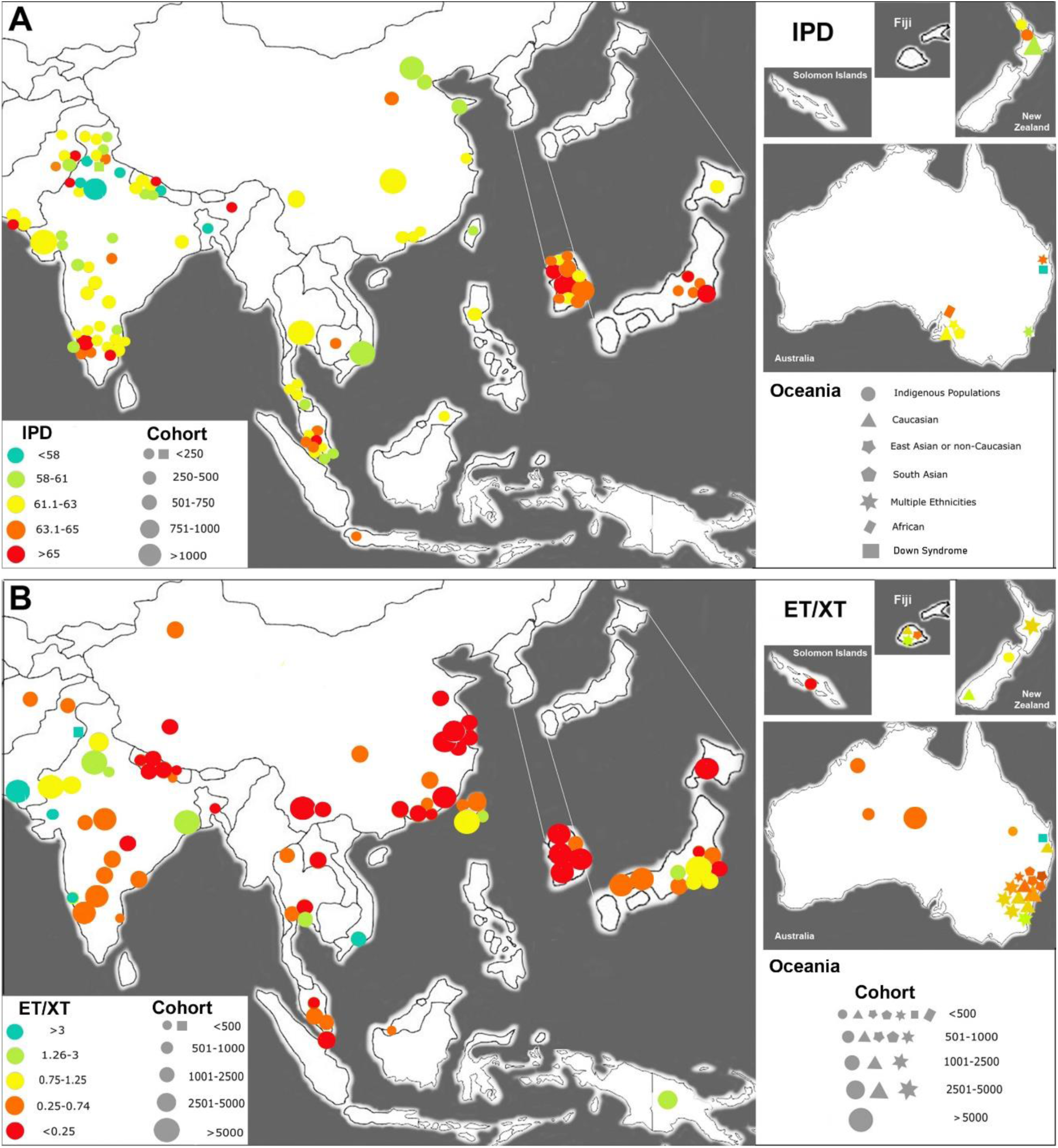
Asia and Oceania. Geographical mapping of the interpupillary distance (IPD) (A) and the esotropia/exotropia ratio (ET/XT ratio) (B) in Asia and Oceania. The size of the circles denotes the size of the cohort examined, and the ranges of the IPDs and the ET/XT ratios are color-coded in a heat map as indicated. Data from Korea and Japan are shown in the magnified regions to better resolve the location of the cohorts. The multiple ethnicities in Oceania are indicated with different symbols, as indicated.

### 2. Pooled estimates of the IPD and subgroup meta-analysis by ethnicity

The pooled mean estimate of the IPD in different ethnicities ranged from 56.1 mm in Down syndrome populations to 69.9 mm in West Africans (Fig. 6A), with Down syndrome = 54.2 to 58.8 mm, Eskimo/Inuit = 59.5 mm, Northwestern South Asia = 60.93 mm, Maoris =61.2 mm, Caucasians (from Europe = 62.3 mm, from Americas, Oceania = 62.5 mm); Middle East = 62.5 mm, East Asians = 62.8 mm, South Asians in South/Central/East = 63.3 mm, Northeast African = 63.9 mm, Latinos/Hispanics = 64.0 mm, Native Americans = 65.1 mm, African Americans = 66.4 mm; West Africans = 69.9 mm. Males had a mean IPD that was 2.0 to 3.3 mm larger than that of females in that same ethnic population, with the exception of people with Down syndrome, where the mean male IPD was 0.9 mm smaller than the mean female IPD (Supplemental Table S1). To calculate a pooled estimate of the IPD that incorporated between-study variation, the pooled IPD means for the major ethnicities were examined by random effects meta-analysis (77% of cohorts qualified for this analysis). Nearly all pooled IPD estimates derived from the complete cohorts were within the 95% confidence intervals defined by the meta-analysis. Two ethnicities were borderline (West Africa at 69.8 mm vs. 69.9 mm; Middle East at 62.7 vs. 62.5 mm), and North/East/South Africa was much lower (63.9 mm) than the low range of the 95% CI, at 66.7 mm, presumably because two studies with nearly half of the total cohort was excluded from the meta-analysis due to lack of SD information. Overall, the meta-analysis supported the mean IPD data reported above and illustrated in Fig. 6A, while controlling for differences in study sample sizes and variability and estimating 95% confidence intervals for point estimates.

### 3. Mapping of horizontal strabismus patterns – esotropia/exotropia (ET/XT) ratios

We report on the global distribution of ET and XT (n=301 studies reporting on 334 cohorts with 2,159,669 subjects).^22^

a. Europe. We found 83 population-based studies that met our inclusion criteria, reporting on ET and XT of 1,090,036 subjects in 88 cohorts (References S1-83, Supplemental Table S3). The highest ET/XT ratio (>3.0) was found in Western and Central Europe, with somewhat lower ET/XT ratios (1.25-3.0; 0.75-1.25) in the northern parts of Sweden, in Finland, Northern parts of Russia, in Portugal, and the Balkans (Fig. 2B). We included ratios reported for people with Down syndrome from four regions in Europe (total of 2,770 subjects).^45^
b. Africa. We found 32 population-based studies reporting on ET and XT (50,100 subjects in 32 cohorts, References S84-115, Supplemental Table S3). Africa has two main types of strabismus patterns. Most populations in West Africa have a low ET/XT ratio (<0.75), while most populations in North Africa (Arabs) and in East Africa (Sudan, Kenya) have more ET than XT with ET/XT ratios between 1.0 and 7.0 (Supplemental Table S2; Fig. 3B).
c. Middle East. We found 32 population-based studies reporting on ET and XT in 154,934 subjects from 33 cohorts (References S116-147, Supplemental Table S3). Most populations in the Middle East have a higher ET/XT ratio (1.25-3.0), with the exception of the population in current Iran, Azerbaijan, and possibly Afghanistan. Nearly all studies (11/14) from Iran report a low ET/XT ratio (0.14-0.77). (Fig. 4B). The large majority of studies from countries surrounding Iran report an ET/XT ratio between 1.48 and 5.0 (Fig. 3D; Supplemental Table S2).
d. Asia, including Oceania. We found a total of 96 population-based studies reporting on 115 cohorts with a total of 636,658 subjects. These included 55 studies reporting on ET and XT in East Asia (total number of subjects in 59 cohorts = 392,141), 25 studies from South Asia (total number of subjects = 131,847 in 25 cohorts), and 18 studies from Oceania (total number of subjects in 31 cohorts = 112,670). (References S148-246, Supplemental Table S3). The lowest ET/XT ratios (<0.25) were reported in East Asia, including China, Nepal, South Korea and Japan. On the Indian subcontinent, most of the populations in the Northwest showed larger ET/XT ratios (1.14-6.25), while nearly all populations in the Central, Southern and Eastern regions had lower ET/XT ratios (0.11-0.69). Populations in Thailand, Malaysia and Singapore also had a lower ET/XT ratio. In Australia, New Zealand and other parts of Oceania, most indigenous populations had lower ET/XT ratios (0.09-1.00), while Caucasians had similar ratios as in North America (1.09-3.33), and populations of East Asian ancestry had lower ET/XT ratios, similar to those in East Asia (0.22-0.63, Fig. 4B).
e. Americas. We found 54 population-based studies reporting on ET and XT, with 35 studies from North America (a total of 197,409 subjects in 45 cohorts), and 19 studies from South and Central America with 54,407 subjects in 21 cohorts (References S247-301, Supplemental Table S3). As detailed for five major ethnicities, ET/XT ratios depended on ethnicity. Nearly all studies on Native Americans report a low ET/XT ratio (0.07-1.00), based on eleven studies from Alaska to the Amazon (Fig. 5B). On the other hand, Inuit (Eskimos) have a high ET/XT ratio of 1.67 to 3.00. Caucasians in the Americas have an ET/XT ratio of 2.25 (n=17 studies in North America, n=8 in South America) that is lower than for Caucasians in Europe (4.35). African Americans (n=8 studies) have an ET/XT ratio of 1.71 (range from 0.22 to 9.5). Latinos/Hispanics in the Americas (n=11 studies) have an ET/XT ratio of 0.92 (range from 0.24-2.00). People with Down syndrome have a high ET/XT ratio (40.43).

**Figure 5:**
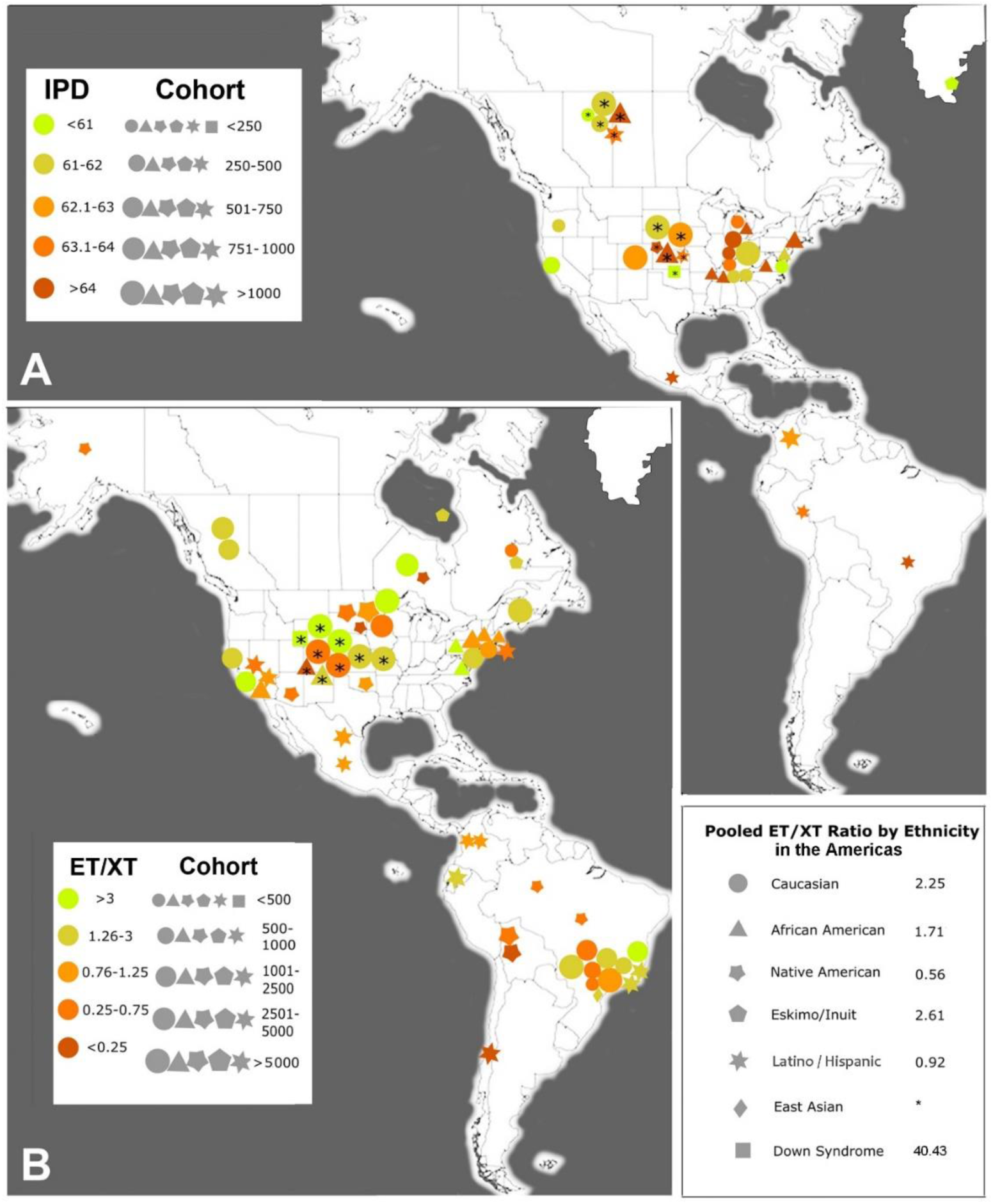
Americas. Geographical mapping of the interpupillary distance (IPD) (A) and the esotropia/exotropia ratio (ET/XT ratio) (B) in the Americas. The size of the symbols denotes the size of the cohort examined. The ranges of the IPDs and ET/XT ratios are color-coded in a heat map as indicated. The multiple ethnicities in North and South America are indicated with different symbols, as indicated in the lower right-hand corner, along with the pooled estimate of the ET/XT ratio for five different ethnicities. *, only one study reported the ET/XT ratio for East Asians within the Americas. Greenland is included in this map because of its vicinity to North America.

### 4. Regression analysis

We compiled the information about the mean IPD (Fig. 6A) and mean ET/XT ratio for all major ethnicities as well for distinct geographic regions (“higher resolution”) and performed regression analyses to determine whether there was a correlation between the two parameters (Supplemental Table S4). Because the ET/XT ratios extend over nearly three orders of magnitude, we used a logarithmic scale for the y-axis (Fig. 6B,C). For the major ethnicities, the trendline was R^2^ = 0.320 between the ET/XT ratio and the IPD (p=0.012041), while for the “higher resolution” regional pairing of values, the trendline was R^2^ = 0.410 (p=0.00000169). This difference likely reflects better matching due to regional pairing, because it takes into account trends and gradients within larger countries. The R^2^ value of 0.410 suggests that 41% of the variation in the ET/XT ratio can be explained by the IPD.

**Figure 6:**
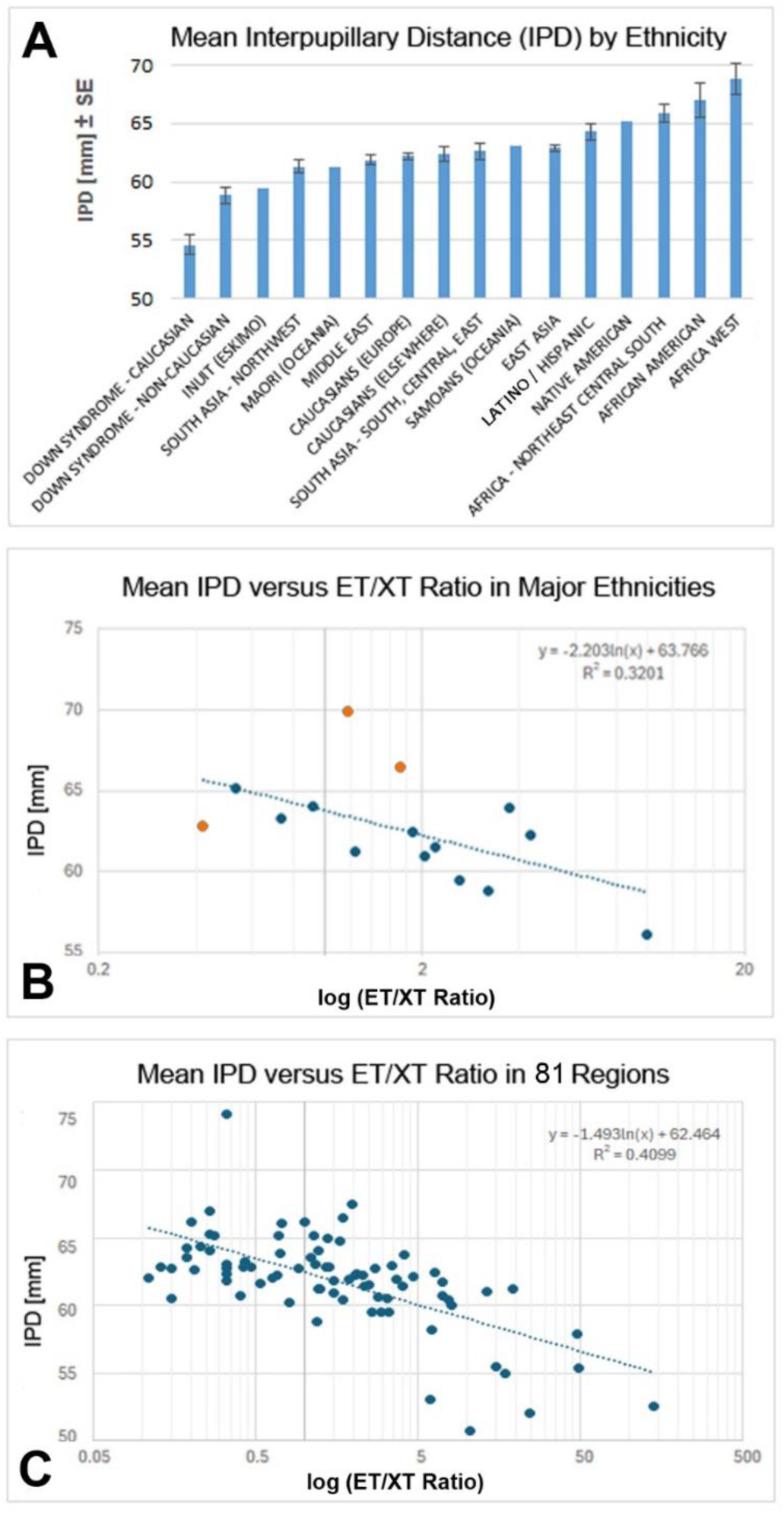
Mean IPD by ethnicity (A) and Regression Analysis of IPD versus ET/XT Ratio for major ethnicities (B) and for regional pairs at higher resolution (C). **A** The mean IPD (average of male and female) for major ethnicities and populations. Error bars show the standard error (SE) when n=2 or more cohorts. The mean IPD for Down syndrome is shown separately for Caucasians (C) and for non-Caucasians (NC). **B** Regression analysis of the mean IPD versus the log of the esotropia/exotropia (ET/XT) ratio for the major ethnicities and populations. The R^2^ value of 0.32 is indicated (p=0.012041). The three orange datapoints are the one farthest from the trendline; the possible significance of these “outliers” is explained in the Results/Discussion. **C** Regression analysis of the mean IPD versus the log (ET/XT ratio) at a higher resolution for 81 different regions where information for both parameters was available (Supplemental Table S4). Note that the R^2^ value increased to 0.41 (p=0.00000169), indicating that 41% of the ET/XT variation may be explained by the IPD.

## Discussion

Our analysis provides the first mapping of the interpupillary distance (IPD) on a regional as well as a global scale. A comprehensive IPD mapping of populations throughout the world has not been previously attempted. Based on our analyses of data from nearly 126,000 subjects, we show that some ethnicities differ substantially in their mean IPDs (Fig. 6A), confirming and significantly extending previous conclusions from a much smaller cohort (3,976 subjects).^4^ Our mapping of the mean IPD may assist the design of proper spectacles for diverse populations, and may also aid in optimizing the range of IPD settings on head-mounted displays for virtual reality applications. This may ensure that adequate ranges of IPDs are covered, and prevent cybersickness due to mismatches of a subject’s IPD and the IPD range on the head-mounted display for virtual reality.^16,18^ The IPD was previously examined in the context of stereoacuity,^213,269^ intelligence (“borderline” significance),^270^ and beauty,^271–272^ and a wider IPD was associated with an extroverted personality.^273^ In our discussion, we focus on the role of the IPD in spectacle design, head-mounted displays, and the association with horizontal strabismus. Our maps show that the mean IPD and the strabismus types (ET/XT ratio) vary between populations, in intriguingly distinct and correlated patterns. Differences in the IPD between ethnicities point to genetic (anatomical) differences that evolved from ancestral orbit types.^274–275^ We explored whether one orbital parameter, the orbital width – as measured by the IPD – is associated with the type of horizontal strabismus as quantified by the ET/XT ratio. Our regression analysis shows that there is a moderate association between the ET/XT ratio and the IPD. We conclude that the orbital anatomy and specifically the IPD contributes to the variation of strabismus patterns, as proposed in the older strabismus literature.^19–20,23–31,276^ In fact, the “growing out of strabismus” for children with esotropia has been explained as being due to the natural growth of the orbit in teenage years.^20,25,277–279^

Why and how does the width of the orbit affect strabismus? Mechanistic models postulate that when the IPD is large, then the medial rectus muscles have to work harder to converge the eyes and achieve binocular fusion, and any failure will tend to produce more XT than ET.^20,23,26,66,276,279^ When IPDs are extreme, either on the narrow side, or on the wide side, then the horizontal muscles are more likely to fail to provide normal alignment, resulting in tropia. This is consistent with the effects of extremely wide (pathological) IPDs in syndromic craniosynostosis such as Apert and Crouzon syndromes, which frequently associate with XT,^278–282^ and the effects of extremely narrow IPDs, as in Down syndrome, which most frequently associate with ET.^45^ The ethnicity-specific distribution of ET/XT ratios in cerebral palsy^283^ are consistent with the proposed effect of the orbital width.

Our regression analysis provides strong evidence that the orbital anatomy plays a role in determining the predominant type of horizontal strabismus. Thus, the higher risk or tendency in certain populations towards either ET or XT is largely genetically determined. The R^2^ values of 0.320 for major ethnicities and 0.410 for populations at higher resolution show an association between the IPD and the ET/XT ratio, indicating that the IPD explains about 41% of the ethnic variation in the ET/XT ratio.

When we examined the paired data points in Figs. 6B and 6C, we found that three ethnicities (West Africans, African Americans, and East Asians) have mean ET/XT ratios that are farthest removed from the global trend line. Values for Africans fall above the trend line, while values for East Asians fall below the trendline. Interestingly, Africans and African Americans have high values of proptosis (protrusion of the eyeball) – another parameter of the orbit besides the IPD which is known to affect horizontal strabismus and varies between ethnicities.^3,284^ Africans have proptosis of about 17 mm,^284–285^ while most East Asian cohorts have values at or below 14 mm.^286–289^ Therefore, the “flat” orbit in East Asians may contribute to the low ET/XT ratio, besides the large IPD, while in Africans, the high proptosis may partially offset the effect of a large IPD, resulting in a higher ET/XT ratio than would be expected based on the IPD alone. Indeed, when the three outliers in Fig. 6B are removed, the R^2^ increases to 0.536, and when the three values are adjusted to lie close to the trendline, the R^2^ reaches 0.761 (data not shown). We propose that the combination of a flat orbit with a wide IPD will cause more XT, while a steep orbit (large proptosis) with a narrow IPD will cause more ET. Additional parameters of the orbit besides IPD and proptosis may further contribute to ethnic differences in strabismus. These include differences in the orbital angle (nasomalar angle), and differences in the insertion or length of the extraocular muscles.^33–34,279,290–295^

Regarding gender differences, we confirm that males generally have a slightly larger IPD than females from the same ethnicity, with the mean gender difference ranging from 2.0 to 3.3 mm between ethnicities (Supplemental Table S1). If a wider IPD associates with more XT, and a narrow IPD with more ET, why do females with a slightly narrower mean IPD not have more ET, and males with their slightly larger mean IPD not have more XT?^296^ It has to be considered that the mean IPD is less relevant than the percentage of subjects in a cohort with extreme IPDs (low or high), since those are most at risk for ET or XT, respectively.^30^ Therefore, the kurtosis of the IPD data is crucial. Kurtosis is rarely reported in studies of the IPD, but in a few studies, the kurtosis can be estimated from the graphs, tables or scatter plots.^4,64,66,96,121,190,235^ Indeed, there is some support from such studies that the kurtosis of the IPD differs between males and females, and this may explain why there is no significant difference between males and females in ET and XT prevalence, despite most studies reporting a gender difference in the mean IPD. Another possibility is a gender difference in the strength (thickness) of some or all the extraocular muscles.^297–298^

The differences in orbital width between populations raises the question of how these differences may have evolved. One plausible and intriguing explanation is that anatomically modern humans encountered and interbred to different extents with archaic hominins such as Neanderthals and Denisovans.^299–304^ Archaic hominins had much wider orbital widths than anatomically modern humans.^304–306^ Depending on the extent of introgression of genes that regulate the width of the developing orbit,^307–308^ populations in East and Central Asia appear to have evolved an orbit that is more disposed to a larger IPD and an eyeball position in the orbit that favors XT over ET. This hypothesis is consistent with the multiple Neanderthal introgression events in East Asia and the geographical distribution of Denisovans.^299,301–303,309^ The ethnic differences then further expanded and became refined by subsequent migrations, 60k to 10k years ago to Australia, and to the Americas as well as within Africa.^309–312^ The wider orbit in present-day West Africans may reflect the fact that they harbor significant amounts of gene introgression from an archaic population that likely diverged before the split of anatomically modern humans and the ancestors of Neanderthals and Denisovans.^312^ Our finding of gradients within India is supported by a recent study showing that populations in the East of India have more Denisovan DNA than populations in the North of India.^313^

Limitations of our review include that not all cohorts are ethnically “pure” or homogeneous – some noise is inevitable. In fact, there has been substantial mixing of populations, especially due to the migrations within Africa, migrations from Europe to North and South America, and within Oceania. Consequences of some of these migrations on the IPD are apparent, for example the Bantu migration from West Africa to Central and South Africa, ^314^ the migration of ancestors of Native Americans from Central Asia, ^315^ the migration from Europe into the Americas, and the migration from Central Asia into Nepal. We acknowledge the lack of data or sparse information for several populations (Native Americans, Inuit/Eskimos, indigenous populations in Oceania). Nevertheless, the narrow IPD in Eskimos is supported by studies examining the interorbital width of skulls, ^316–318^ and likewise the relatively large interorbital width in Native Americans. ^319^ Ideally, we would like to have information about the IPD and the ET/XT ratio from exactly the same cohorts, but such studies are sparse.^54,56,85–86,103,238,246^ The rarely reported kurtosis may be more relevant as a risk factor for strabismus than the mean IPD.

## Conclusion

The interpupillary distance differs between ethnicities and associates with horizontal strabismus patterns. A narrow orbit is associated with an increased frequency of esotropia, and a wider orbit is associated with an increased frequency of exotropia. The significance of our work is that it provides a better understanding of horizontal strabismus, its development and evolution, and greater appreciation for diverse strabismus patterns; this is important for planning of vision care and risk factor awareness. Our work may also help with appropriate spectacle design and proper head-mounted displays that are becoming increasingly popular in virtual reality.^320^

## Data Availability

All data produced in the present work are contained in the manuscript

## Acknowledgments

The authors thank the following individuals who helped to obtain relevant literature and/or assisted with translations of texts: Aderonke Baiyeroju (Univ. of Ibidan, Nigeria), Jenny Costa (University of Nevada, Reno), Chengyuan Feng (Johns Hopkins University, USA), Koji Matsuda (Matsuda Eye Clinic, Osaka, Japan), Sineenart Sengyee (University of Nevada, Reno), Napaporn Tananuvat (Chiang Mai University, Thailand), Carina Vetye-Maler (Munich/Buenos Aires), and Wei Yang (University of Nevada, Reno). Grant support: NIH grants EY031729 (C.S.v.B.) and GM103554 (C.S.v.B.), International Research Support Initiative Program (IRSIP) fellowship by the Higher Education Commission of Pakistan (Z.Z.).

## SUPPLEMENTAL TABLES

**Supp. Table 1.** Reports of the interpupillary distance (IPD) in different populations/regions. The mean difference between males (M) and females (F) for each ethnicity with data is indicated as: M=F+x (mm). Reported values for the IPD are shown in bold font and estimated/calculated values (for male, female or the mean) are shown in regular font. Ref, reference showing the number in the List of References.

| First Author | Ref # | Year | Ethnicity/Region | Comments | Cohort size | Male | Female | Mean |
| --- | --- | --- | --- | --- | --- | --- | --- | --- |
| **EUROPE** | | | | $M=F+2.3$ | | | | |
| Becker | 58 | 1861 | Heidelberg, Germany | c/o Beselin | 370 | **60.1** | **57.5** | 58.8 |
| Pflüger | 59 | 1875 | Luzerne, Switzerland |  | 214 | **62.0** | **59.2** | 61.4 |
| Holmgren | 60 | 1879 | Upsala, Sweden |  | 191 | **62.6** | **61.5** | 62.1 |
| Beselin | 61 | 1885 | Heidelberg, Germany |  | 8 | 63.1 | **60.8** | 61.95 |
| Seggel | 62 | 1904 | German soldiers |  | 5,000 | **62.2** | 59.8 | 61.0 |
| Seggel | 62 | 1904 | Munich, Germany |  | 700 | **62.5** | **61.5** | 62.0 |
| Speidel | 63 | 1905 | Tubingen, Germany |  | 472 | **62.4** | 60.1 | 61.25 |
| Helmbold | 64 | 1914 | Germany |  | 300 | **65.2** | **62.1** | 63.7 |
| Helmbold | 64 | 1914 | Eastern Germany |  | 225 | **64.1** | **61.1** | 62.6 |
| Koegel | 65 | 1914<br>1916 | Belarus |  | 296 | **65.2** | 62.9 | 64.05 |
| Koegel | 66 | 1914 | East-Central Russia |  | 300 | **64.5** | 62.2 | 63.35 |
| Koegel | 66 | 1914 | Ukraine |  | 300 | **64.9** | 62.6 | 63.75 |
| Koegel | 66 | 1914 | Tatarstan, Russia | Tatars | 300 | **63.7** | 61.4 | 62.55 |
| Guenther | 1 | 1933 | Saxony, Germany |  | 100 | **65.8** | 63.5 | 64.65 |
| Bertelsen | 67 | 1954 | Copenhagen, Denmark |  | 249 | **62.06** | **59.85** | **60.81** |
| Kerwood | 68 | 1954 | Eastern England, UK |  | 200 | **64.3** | **60.6** | 62.45 |
| Babalola | 69 | 1960 | UK |  | 40 | **64.7** | 62.4 | 63.55 |
| Kisling | 70 | 1966 | Copenhagen, Denmark |  | 102 | **61.0** | 58.7 | 59.85 |
| Garrett | 71 | 1971 | France | Pilots | 820 | **62.3** | 60.0 | 61.15 |
| Bruckner | 72 | 1986 | Basel, Switzerland |  | 11 | **65.16** | 62.86 | 62.83 |
| Fledelius | 37 | 1986 | Hillerod, Denmark |  | 203 | **65.9** | **62.9** | 64.4 |
| Jonasson | 73 | 1987 | Iceland |  | 555 | 61.85 | 59.55 | **60.7** |
| Mon-Williams | 12 | 1993 | Scotland |  | 20 | 62.15 | 59.85 | **61.0** |
| Howarth | 74 | 1999 | Leicestershire, UK |  | 41 | **64.0** | **61.0** | 62.5 |
| Pointer | 75 | 1999 | Northampton, UK |  | 900 | **64.8** | **61.7** | 63.3 |
| Pointer | 75 | 1999 | Northampton, UK |  | 900 | **65.5** | **62.6** | 64.1 |
| Filipovic | 76 | 2003 | Croatia |  | 200 | 64.15 | 61.85 | **63.0** |
| Wilkinson | 77 | 2003 | Manchester, UK |  | 95 | **60.8** | **58.5** | 59.7 |
| Mommaerts | 78 | 2008 | Belgium |  | 50 | **60.6** | **59.4** | 60.0 |
| Pointer | 9 | 2012 | Northampton, UK |  | 1,354 | **65.3** | **62.3** | 63.8 |
| Garcia-Lazaro | 79 | 2014 | Valencia, Spain |  | 38 | 64.3 | 61.9 | **63.1** |
| Lupon-Bas | 80 | 2014 | Barcelona, Spain |  | 5 | 63.15 | 60.85 | **62.0** |
| Zilch | 13 | 2014 | Hamburg, Germany |  | 15 | 67.25 | 64.95 | **66.1** |
| Hakala | 81 | 2016 | Espoo, Finland |  | 40 | 61.55 | 59.25 | **60.4** |
| Mestre | 14 | 2016 | Marseille, France |  | 20 | **64.1** | 61.78 | 62.95 |
| Flament | 82 | 2020 | Paris, Montpellier, France |  | 581 | 65.3 | **63.0** | 64.15 |
| Knezi | 83 | 2020 | Serbia |  | 90 | **60.3** | **60.8** | 60.5 |
| Schuetz | 17 | 2022 | Giessen, Germany |  | 18 | 62.25 | 59.95 | **61.1** |
| Tsiogka | 84 | 2023 | Athens, Greece |  | 400 | 62.15 | 59.85 | **61.0** |
| DOWN SYNDROME |  |  |  | M=F-2.4 |  |  |  |  |
| Brushfield | 85 | 1924 | London, UK |  | 84 | **50** | **54** | 52 |
| Vontobel | 54 | 1933 | Zurich, Switzerland |  | 10 | 49.5 | 51.9 | **50.7** |
| Lowe | 86 | 1949 | London, UK |  | 17 | 53.8 | 56.2 | **55** |
| Kerwood | 68 | 1954 | Eastern England, UK |  | 33 | **54.6** | **55.4** | 55.0 |
| Kisling | 70 | 1966 | Denmark (South) |  | 71 | **54.2** | 56.6 | 55.4 |
| Missiroli | 55 | 1970 | Rome, Italy |  | 15 | **59.2** | 61.6 | 60.4 |
| Woodhouse | 56 | 1994 | Cardiff, Wales |  | 8 | 48.8 | 51.2 | **50** |
| Doyle | 87 | 2016 | Ulster, UK |  | 7 | **53.2** | 55.6 | 54.4 |
| INUIT (ESKIMO) |  |  |  |  |  |  |  |  |
| Skeller | 88 | 1954 | Tasiilaq, S. Greenland |  | 76 | **60.26** | **58.67** | 59.47 |
| AFRICA – WEST |  |  |  |  |  |  |  |  |
|  |  |  |  | M=F+2.27 |  |  |  |  |
| Babalola | 69 | 1960 | West Africa |  | 40 | **69.64** | 67.5 | 68.5 |
| Kikudi | 30 | 1988 | Kinshasa, Zaire |  | 100 | 67.15 | 64.85 | **66** |
| Kaimbo | 89 | 1994 | Kinshasa, Zaire |  | 11 | **70.25** | **65.3** | **67.8** |
| Umar | 90 | 2005 | Jos, Central Nigeria |  | 213 | **89** | 86.7 | 87.85 |
| Ilechie | 91 | 2010 | Cape Coast, Ghana |  | 400 | **69.6** | **66.3** | **68** |
| Oladipo | 92 | 2010 | Bayelsa State, Nigeria | Ijaw | 1,000 | **69.60** | **66.64** | **68.1** |
| Osunwoke | 93 | 2010 | Enugu-Owerri, Nigeria | Igbo | 1,000 | **74.9** | **74.5** | **74.7** |
| Esomonu | 94 | 2012 | Enugu-Owerri, Nigeria | Igbo | 2,000 | **74.25** | **74.2** | 74.2 |
| Osunwoke | 42 | 2012 | Port Harcourt, Nigeria | Ijaw | 175 | **65.08** | **65.01** | 65.05 |
| Kumah | 95 | 2016 | Kumasi, Ghana |  | 302 | **66.2** | **65.2** | **65.7** |
| Omodele | 96 | 2016 | Yoruba, Nigeria |  | 1,274 | **68.8** | **66.58** | **67.7** |
| Usman | 97 | 2016 | North-Eastern Nigeria | Bura | 300 | **69.6** | **68.0** | **69.0** |
| Adekunle | 98 | 2021 | Lagos, Nigeria | Yoruba | 325 | **68.19** | **65.35** | **66.99** |
| Adekunle | 98 | 2021 | Lagos, Nigeria | Igbo | 80 | **68.36** | **66.24** | **67.34** |
| Adekunle | 98 | 2021 | Lagos, Nigeria | Hausa | 50 | **66.86** | **63.41** | **66.73** |
| Adekunle | 98 | 2021 | Lagos, Nigeria | other | 48 | 68.23 | 65.96 | **67.09** |
| Adekunle | 99 | 2022 | Lagos, Nigeria |  | 452 | **69.63** | **65.54** | **67.85** |
| Soumboundou | 100 | 2023 | Dakar, Senegal |  | 104 | **61.9** | **59.9** | 60.9 |
| DOWN SYNDROME |  |  |  |  |  |  |  |  |
| Adio | 101 | 2012 | Port Harcourt, Nigeria |  | 42 |  |  | **59.5** |
| AFRICA – NORTHEAST, CENTRAL, SOUTH |  |  |  | M=F+2.4 |  |  |  |  |
| Ried | 102 | 1915 | Congo, Tanganyika |  | 37 | **68** | **63.3** | 65.65 |
| El-Mokadem | 103 | 1987 | Egypt |  | 2,600 | **63.4** | **60** | **61.7** |
| Rasengane | 104 | 2001 | Johannesburg, South Africa |  | 200 | 68.1 | 65.9 | **67** |
| McAlister | 105 | 2002 | Uganda (Army) |  | 104 | **67.4** | 65.0 | 66.2 |
| Ruiz-Alcocer | 106 | 2011 | Maputo, Mozambique |  | 423 | **68.1** | **67.4** | **67.8** |
| Siraj | 107 | 2017 | Khartoum, Sudan |  | 920 | **63.0** | **61.57** | **62.3** |
| Halladay | 108 | 2019 | Malawi |  | 179 | **68.6** | **65.6** | **67.8** |
| Elrazky | 109 | 2020 | Cairo, Egypt |  | 83 | 65.7 | 61.3 | **63.5** |
| Flament | 82 | 2020 | Johannesburg, South Africa |  | 315 | 71.1 | **68.7** | 69.9 |
| Mhaleni | 110 | 2021 | Polokwane, Limpopo Province, South Africa |  | 386 | **68** | **66.5** | **67.3** |
| Samuel | 111 | 2021 | Mashonaland, Zimbabwe | Shona | 471 | **66.4** | **65.0** | **65.7** |
| **MIDDLE EAST** |  |  |  | M=F+2.44 |  |  |  |  |
| Osuobeni | 112 | 1993 | Riyadh, Saudi Arabia |  | 353 | 63.4 | **60.9** | 62.15 |
| Osuobeni | 113 | 1994 | Riyadh, Saudi Arabia |  | 198 | **63.38** | 60.94 | 62.16 |
| Al-El-Sheikh | 114 | 1998 | Riyadh, Saudi Arabia |  | 163 | **64.9** | **59.4** | **62.15** |
| Evereklioglu | 115 | 1999 | Malatya, Turkey |  | 1,404 | **63.7** | **61.5** | **62.6** |
| Evereklioglu | 116 | 2002 | Malatya, Turkey |  | 1,414 | **63.7** | **61.5** | **62.6** |
| Bozkir | 117 | 2003 | Adana, Turkey |  | 1,404 | **63.9** | **60.8** | **62.3** |
| Kassab | 118 | 2005 | Mosul, Iraq |  | 100 | **64.9** | **59.4** | 62.15 |
| Bayani | 119 | 2006 | Tehran, Iran |  | 400 | **65** | **63** | **64** |
| Ozturk | 120 | 2006 | Afyon, Turkey (West) |  | 261 | **61.4** | **61.0** | **61.2** |
| Beden | 121 | 2008 | Samsun, Turkey |  | 2,477 | 64.2 | 61.76 | **62.98** |
| Ozsoy | 122 | 2009 | Antalya, Turkey |  | 70 | 64.25 | 61.75 | **63.0** |
| Aksu | 123 | 2010 | Ankara, Turkey |  | 100 | **48.3** | **51.4** | **49.9** |
| Mahdaviizadi | 124 | 2010 | Mashhad, Iran |  | 100 | 63.54 | 60.88 | **62.1** |
| Fesharaki | 125 | 2012 | Isfahan, Iran |  | 947 | **63.6** | **61.1** | **62.4** |
| Alanazi | 126 | 2013 | Riyadh, Saudi Arabia |  | 133 | **62.65** |  | 61.40 |
| Aslankurt | 127 | 2013 | Kahramanmaras, Turkey |  | 120 | **64.4** | **61.3** | 62.85 |
| Gudek | 128 | 2015 | Samsun, Turkey |  | 115 | **61.7** | **59.0** | **60.4** |
| Ilhan | 10 | 2015 | Kayseri, Turkey |  | 1,002 | 61.12 | 58.68 | **59.9** |
| Yildirim | 129 | 2015 | Istanbul, Turkey |  | 756 | **63.9** | **61.4** | **62.7** |
| Al-Kaisy | 130 | 2016 | Kurds in Iraq |  | 65 | 59.82 | 57.38 | **58.6** |
| Direk | 131 | 2016 | Sanliurfa, Turkey |  | 311 | 66.64 | **64.2** | 65.42 |
| Moravej | 132 | 2017 | Tehran (W), Iran |  | 487 | 61.44 | **59** | 60.22 |
| Ozdemir | 133 | 2017 | Bursa, Turkey |  | 172 | **59.3** | **57.5** | **58.4** |
| Majeed | 134 | 2018 | Abha, Saudi Arabia |  | 228 | **65.87** | **57.79** | **61.9** |
| Rabaji | 135 | 2019 | Tehran, Iran |  | 24 | **66.03** | **64.60** | 65.32 |
| Sahbaz | 136 | 2020 | Azeri population |  | 1,132 | **65** | **62.7** | **63.9** |
| Sahbaz | 137 | 2021 | Azerbaijan |  | 700 | **66.0** | **64.1** | **65.05** |
| Bahsi | 138 | 2021 | Gaziantep, Turkey |  | 200 | **67.6** | **63.2** | **65.4** |
| Gantz | 139 | 2021 | Jerusalem, Israel | Jews, Arabs | 199 | **61.9** | **60.0** | **60.6** |
| Sahbaz | 137 | 2021 | Istanbul, Turkey |  | 700 | **63.1** | **61.1** | **62.1** |
| Moradi | 140 | 2022 | Tehran, Iran |  | 300 | **63.26** | **61.13** | **61.95** |
| Ozdemir | 141 | 2022 | Samsun, Turkey |  | 80 | **66.15** | **65.21** | 65.68 |
| Alshamri | 142 | 2023 | Yemen |  | 150 | **60.8** | **59.9** | 60.35 |
| Hatipoglu | 143 | 2023 | Nigde, Turkey |  | 57 | **64** | **62** | **63** |
| Abbas | 144 | 2024 | Najaf, Iraq |  | 77 | 63.6 | 61.16 | **62.38** |
| Eltahir | 145 | 2024 | AL-Qassim, Saudi Arabia |  | 56 | 60.53 | **58.09** | 59.31 |
| **SOUTH ASIA (NORTHWEST)** |  |  |  | M=F+2.68 |  |  |  |  |
| Singh JR | 146 | 1983 | Amritsar, Punjab, India |  | 71 | **59.1** | **56.5** | 57.8 |
| Gupta VP | 147 | 2003 | Delhi, India |  | 1,516 | **58.5** | **57** | 57.8 |
| Bali | 148 | 2005 | Kinnaur/Lahaul-Spiti, India |  | 50 | 66.4 | 63.72 | **65.06** |
| Patil | 149 | 2011 | Nagpur, India |  | 216 | **64.2** | **63.1** | 63.7 |
| Hussain | 150 | 2012 | Lahore, Pakistan |  | 159 | **67.57** | **62.92** | 65.2 |
| Kini | 151 | 2012 | Loni, Maharashtra, India |  | 70 | 63.34 | 60.66 | **62** |
| Sharma | 152 | 2012 | Sundar Nagar, India |  | 100 | **59.8** | **57.6** | 58.7 |
| Purkait R | 153 | 2013 | Sagar, India |  | 116 | **60.7** | **57.4** | 59.1 |
| Ladda | 154 | 2014 | Loni, Maharashtra, India |  | 400 | **61.1** | **58.2** | 59.65 |
| Shah | 43 | 2014 | Gujarat, India |  | 1,696 | **61.5** | **60** | **61.25** |
| Nazir | 155 | 2015 | Srinagar, Kashmir, India |  | 60 | **63.5** | **61.9** | **62.0** |
| Alkhairy | 156 | 2016 | Karachi, Pakistan |  | 500 | **61.9** | **61.5** | 61.7 |
| Gupta R | 157 | 2016 | Shimla, Himachal Pradesh, India |  | 40 | **67.9** | **61** | 64.5 |
| Arshad | 158 | 2017 | Faisalabad, Pakistan |  | 100 | 65.34 | 62.66 | **64.0** |
| Ayoub | 159 | 2017 | Kashmir, India |  | 60 | **60.2** | **59.5** | 59.85 |
| Bangar | 160 | 2017 | Latur, Maharashtra, India |  | 250 | **62.3** | **61.5** | 61.9 |
| Batool | 161 | 2017 | Faisalabad, Pakistan |  | 30 | 63.4 | 60.4 | **62.6** |
| Shah DS | 162 | 2017 | Gandhinagar, Gujarat, India |  | 100 | **60.3** | **58.79** | 59.55 |
| Saifullah | 163 | 2018 | Peshawar, Pakistan |  | 100 | **63.7** | **61.0** | **62.3** |
| Hayat | 164 | 2019 | Karachi, Pakistan |  | 499 | **62.7** | **60.7** | 61.7 |
| Rattan | 165 | 2019 | Uttarakhand, India |  | 100 | **59.4** | **48.3** | 53.9 |
| Gupta P | 166 | 2020 | Bathinda, Punjab, India |  | 200 | **67.2** | **65.9** | 66.5 |
| Dhinsa | 167 | 2021 | Haryana, India |  | 100 | **54.6** | **51.6** | 53.1 |
| Sarathi | 168 | 2021 | Gujarat+, India |  | 71 | 63.84 | 61.16 | **62.5** |
| Bhalla | 169 | 2022 | Chandigarh, India |  | 45 | **61.9** | **60.8** | 61.35 |
| Ahmed | 170 | 2022 | Karachi, Pakistan |  | 230 | **66.62** | **71.72** | **69.23** |
| Malik | 171 | 2022 | Lahore, Pakistan |  | 380 | **60.23** | **58.86** | **59.6** |
| Poonia | 172 | 2022 | Ahmedabad, India |  | 150 | 60.85 | 58.15 | **59.5** |
| Kamboj | 173 | 2023 | Haryana, India |  | 100 | **62.6** | **61.55** | **62.05** |
| Maseedupalli | 11 | 2023 | Hyderabad, India |  | 455 | **62.8** | **60.0** | 61.4 |
| DOWN SYNDROME |  |  |  |  |  |  |  |  |
| Bhalla | 57 | 2021 | Chandigarh, India |  | 51 | **58.3** | **58.0** | 58.15 |
| **SOUTH ASIA (SOUTH, CENTRAL, EAST)** |  |  |  | M=F+3.07 |  |  |  |  |
| Anitha | 174 | 2011 | Chennai, India |  | 100 | **64** | **58** | 61 |
| Packiriswamy | 175 | 2012 | Manipal, Karnataka, India |  | 300 | **68.1** | **63.6** | 65.9 |
| Vasanthakumar | 176 | 2013 | Manipal, Karnataka, India | Dravidians | 200 | **66.72** | **62.59** | 64.7 |
| Mostafa | 177 | 2014 | Chittagong/Rangamati, Bangladesh | Buddhist | 100 | 56.9 | **55.3** | 56.1 |
| Shivhare | 178 | 2015 | Bangalore, India |  | 90 | **63.1** | **60.6** | 61.9 |
| Narain | 179 | 2016 | Davanagere, Karnataka, India |  | 200 | **63.1** | **60.6** | **63.01** |
| Banu | 180 | 2017 | Mangalore, Karnataka |  | 120 | **65.2** | **61.8** | 63.5 |
| Barman | 181 | 2018 | Meghalaya, Assam, India |  | 120 | **70.86** | **70.0** | 70.43 |
| Sasidharan | 182 | 2019 | Mangaluru, Karnataka |  | 400 | **62.1** | **61.3** | **61.7** |
| Sivaranjani | 183 | 2019 | Chennai, Tamil Nadu |  | 100 | **62** | **62** | 62 |
| Yu | 184 | 2019 | Indians residing in Singapore |  | 70 | **62.7** | **57.6** | 60.2 |
| Flament | 82 | 2020 | Chennai, Kolkata, Delhi, India |  | 886 | 66.07 | **63.0** | 64.34 |
| Nehaapriya | 185 | 2020 | Chennai, Tamil Nadu |  | 150 | 60.03 | 56.97 | **58.5** |
| Rupashri | 186 | 2020 | Chennai, Tamil Nadu |  | 250 | **62.7** | **61.8** | 62.3 |
| Deepasakthi | 187 | 2021 | Chennai, Tamil Nadu |  | 102 | **66.91** | **63.39** | 65.15 |
| Sarathi | 168 | 2021 | Tamil Nadu+, India |  | 78 |  |  | 62.9 |
| Shetty | 188 | 2021 | Mangalore, Karnataka, India |  | 252 | **62.26** | **58.46** | 60.36 |
| Girimallanavar | 189 | 2022 | Bangalore, Karnataka, India |  | 100 | **65.1** | **60.9** | 63.0 |
| Singh AD | 190 | 2023 | Andhra Pradesh, India |  | 100 | 64.41 | 61.35 | **62.88** |
| Bandhu | 191 | 2023 | Jharkhand, India |  | 500 | **63.3** | **61** | **62.18** |
| **EAST ASIA** |  |  |  | M=F+2.55 |  |  |  |  |
| Fung | 192 | 1964 | Beijing, China |  | 1,113 | **61.29** | **59.85** | 60.57 |
| White | 193 | 1964 | Thailand (South) |  | 2,950 | **64.0** | 61.5 | 62.75 |
| White | 194 | 1964 | Vietnam |  | 2,129 | **62.0** | 59.5 | 60.75 |
| Pryor | 36 | 1969 | Japanese (in USA) |  | 149 | **66.0** | **64.0** | 65.0 |
| Nakagawa | 195 | 1974 | Sapporo, Japan |  | 281 | **63.5** | **61.6** | 62.6 |
| Park YK | 196 | 1975 | Seoul (South), Korea |  | 570 | **65.31** | **60.78** | 63.05 |
| Sun | 197 | 1979 | Tianjin, China |  | 650 | **61.0** | 58.5 | 59.75 |
| Wang JL | 198 | 1981 | Guanxi Zhuang, China |  | 944 | **62.0** | **60.3** | 61.2 |
| Kim CJ | 199 | 1988 | Korea (cited by Vasanthakumar, 2013) |  | 323 | **59.9** | **63.5** | 61.7 |
| Park | 38 | 1990 | Daegu, Korea |  | 1,653 | **65.45** | **64.30** | 64.88 |
| Quant | 200 | 1992 | Hong Kong, China |  | 243 | **64.59** | **61.31** | 62.95 |
| Quant | 201 | 1992 | Guangdong, China |  | ~200 | **63.72** | **61.04** | 62.38 |
| Cho JH | 202 | 1993 | Korea |  | 1,500 | **69.4** | **66.6** | 68.0 |
| Hwang | 203 | 1996 | Seoul, Korea |  | 700 | **69.4** | **66.6** | **67.6** |
| Tang | 204 | 1998 | Hong Kong, China |  | 500 | **64.3** | **61.6** | 62.95 |
| Song | 205 | 1999 | Daegu, Korea |  | 498 | **64.6** | **63.6** | 64.1 |
| Wang Y | 40 | 2001 | Qingdao+, China |  | 740 | **61.5** | **59.37** | **60.39** |
| Ngeow | 206 | 2006 | Sabah, Malaysia | KadasanDusun | 140 | **64.6** | **61.2** | 62.9 |
| Al Junid | 207 | 2007 | Kuala Lumpur, Malaysia | Malays | 100 | **64.5** | **62.3** | **63.4** |
| Al Junid | 207 | 2007 | Kuala Lumpur, Malaysia | Chinese | 100 | **64.8** | **61.4** | **63.1** |
| Al Junid | 207 | 2007 | Kuala Lumpur, Malaysia | Indians | 100 | **64.1** | **61.1** | **62.6** |
| Du | 208 | 2008 | Hubei, Jianxi, Chongqing, Guangxi, Shanxi (China) |  | 3,000 | **64.2** | **61.1** | 62.6 |
| Lee | 209 | 2008 | Daejeon, Korea |  | 275 | **62.3** | **59.9** | 61.1 |
| Lee | 209 | 2008 | Ueda, Nagano, Japan |  | 275 | **68.9** | **63.9** | 66.4 |
| Park DH | 210 | 2008 | Daegu, Korea |  | 498 | **64.6** | **63.6** | 64.1 |
| Isa | 211 | 2010 | Kuala Lumpur, Malaysia | Malays | 61 | 63.58 | 61.03 | **62.3** |
| Preechawai | 212 | 2011 | Thailand (South) | Chinese | 24 | **64.4** | **59.7** | 62.1 |
| Preechawai | 212 | 2011 | Thailand (South) | Thai | 24 | **61.3** | **59.5** | 60.5 |
| Preechawai | 212 | 2011 | Thailand (South) | Thai-Malay | 24 | **62.9** | **60.7** | 61.8 |
| Preechawai | 212 | 2011 | Thailand (South) | Thai-Chinese | 24 | **63.8** | **60.0** | 61.9 |
| Eom | 213 | 2013 | Seoul, Korea |  | 33 | 64.38 | 61.83 | **63.1** |
| Takahashi | 214 | 2014 | Aichi, Japan |  | 143 | **64.7** | **61.9** | 63.3 |
| Mishra MK | 215 | 2016 | Dharan, Nepal | Mongoloid | 85 | 58.78 | 56.23 | **57.5** |
| Mishra MK | 215 | 2016 | Dharan, Nepal | Aryans | 85 | 63.2 | 60.65 | **61.92** |
| Lin | 216 | 2017 | Taiwan, China |  | 206 | **61.6** | **58.8** | 60.2 |
| Lu | 217 | 2017 | Kuala Lumpur, Malaysia | Malays | 103 | **66.03** | **62.44** | **64.04** |
| Lu | 217 | 2017 | Kuala Lumpur, Malaysia | Chinese | 97 | **64.37** | **61.33** | **62.85** |
| Hwang H | 218 | 2018 | Daejeon, Korea |  | 21 | 65.48 | 62.93 | **64.2** |
| Kim YC | 219 | 2018 | Seoul, Korea |  | 91 | 63.35 | **61.0** | 62.2 |
| Rokaya | 220 | 2018 | Kathmandu, Nepal |  | 200 | **62.67** | **59.46** | 61.07 |
| Mishra S | 221 | 2019 | Bharatpur, Nepal |  | 113 | 62.0 | **59.5** | 60.75 |
| Sano | 222 | 2019 | Tochigi, Japan |  | 213 | 64.38 | 61.83 | **63.1** |
| Yu P | 183 | 2019 | Residing in Singapore | Chinese | 150 | **61.9** | **59.2** | 60.6 |
| Flament | 82 | 2020 | China (multiple sites) |  | 574 | 67.55 | **65.0** | 66.3 |
| Flament | 82 | 2020 | Tokyo, Japan |  | 915 | 68.55 | **66.0** | 67.3 |
| Jung | 223 | 2020 | Daegu, Korea |  | 120 | 64.6 | **62.2** | 63.4 |
| Aziz | 224 | 2021 | Selangor, Malaysia |  | 170 | **68.3** | **65.9** | 67.2 |
| Kim JS | 18 | 2021 | Seoul, Korea |  | 22 | **66.8** | **60.5** | 63.65 |
| Husna | 225 | 2022 | Java (West) |  | 39 | 65.1 | **62.6** | 63.85 |
| Tuladhar | 226 | 2023 | Pokhara, Nepal |  | 199 | **67.13** | **65.15** | **66.09** |
| Xu Y | 227 | 2023 | Shanghai, China |  | 103 | 63.95 | 61.6 | **62.75** |
| Hor | 228 | 2023 | Cambodia |  | 50 | 64.36 | 61.81 | **63.08** |
| Alberto | 229 | 2024 | Cabanatuan City, Philippines |  | 300 | **62.59** | **60.29** | 61.44 |
| Laher | 230 | 2024 | Dang Valley, Nepal | Aryans | 406 | **62.25** | **60.48** | 61.37 |
| Laher | 230 | 2024 | Dang Valley, Nepal | Mongoloids | 406 | **62.71** | **61.91** | 62.31 |
| Wang J | 231 | 2024 | Taiyuan, Shanxi, China |  | 409 | **65.43** | **61.64** | **63.49** |
| **OCEANIA** |  |  |  |  |  |  |  |  |
| INDIGENOUS |  |  |  |  |  |  |  |  |
| Bridgman | 232 | 1999 | New Zealand, Maori |  | 191 | **63.3** | **60.1** | 61.2 |
| Bridgman | 232 | 1999 | New Zealand, Samoan |  | 148 | **64.5** | **61.7** | 63.1 |
| MULTIPLE ETHNICITIES |  |  |  |  |  |  |  |  |
| Swan | 233 | 2005 | Adelaide, Australia | Caucasian | 54 | **63.6** | **59.6** | 61.6 |
| Ellakwa | 234 | 2011 | Sydney, Australia | Multiple Ethnicities | 120 | **62.01** | **58.91** | **60.68** |
| Kolose | 235 | 2021 | New Zealand, Mixed Populations | Defense Force | 1,002 | **61** | **58** | 59.5 |
| Rana | 236 | 2023 | Adelaide, Australia | Caucasian | 316 | 62.7 | 59.7 | **61.2** |
| Rana | 236 | 2023 | Adelaide, Australia | East Asian | 28 | 63.2 | 60.2 | **61.7** |
| Rana | 236 | 2023 | Adelaide, Australia | South Asian | 27 | 63.9 | 59.9 | **61.4** |
| Rana | 236 | 2023 | Adelaide, Australia | African | 9 | 66.8 | 63.8 | **65.3** |
| McAnally | 237 | 2024 | Brisbane, Australia |  | 30 | 65.2 | 62.2 | **63.7** |
| DOWN SYNDROME |  |  |  |  |  |  |  |  |
| Fanning | 238 | 1971 | Brisbane, Australia |  | 24 |  |  | **53.0** |
| **AMERICA** |  |  |  |  |  |  |  |  |
| CAUCASIAN |  |  |  | M=F+2.64 |  |  |  |  |
| Jackson | 35 | 1921 | Colorado, USA |  | 4,560 | **63.7** | **61.5** | 62.6 |
| Davenport | 239 | 1929 | Jamaica |  | 100 | **65.12** | **62.60** | 63.86 |
| Lucas | 240 | 1935 | San Francisco, CA, USA |  | 370 | **59** | **59** | **59** |
| Hertzberg | 241 | 1954 | Ohio, USA |  | 4,057 | **63.27** | 60.6 | 61.95 |
| Ditmars | 29 | 1966 | San Francisco, CA, USA |  | 500 | **65.44** | **62.62** | **64.18** |
| Pryor | 36 | 1969 | San Francisco, CA, USA |  | 391 | **62.0** | 59.35 | 60.65 |
| White | 242 | 1971 | US Army, USA |  | 6,680 | **61.3** | 58.65 | 60.0 |
| Backman | 243 | 1972 | Quebec, Canada |  | 5 | 66.32 | 63.68 | **65.0** |
| Hofstetter | 244 | 1972 | Indiana, USA |  | 331 | **66.5** | 64.0 | 65.3 |
| Hofstetter | 244 | 1972 | Indiana, USA |  | 134 | **65.7** | 63.2 | 64.5 |
| Hofstetter | 244 | 1972 | Indiana, USA |  | 113 | **64.4** | 61.9 | 63.2 |
| White | 245 | 1977 | US Marine Corps, USA |  | 2,008 | **60.8** | 58.1 | 59.45 |
| Jaeger | 246 | 1980 | Philadelphia, PA, USA |  | 88 | **61.4** | **60.0** | 60.7 |
| Cesario | 5 | 1984 | Washington DC, USA |  | 50 | **57.87** | **56.61** | 57.2 |
| Bogren | 247 | 1986 | Sacramento, CA, USA |  | 7 | 61.66 | **59** | **60.3** |
| Latta | 6 | 1991 | Tennessee, USA |  | 54 | **63** | **60** | 61.5 |
| Young | 248 | 1993 | USA |  | 238 | **61.39** | **58.44** | 59.92 |
| Farkas | 249 | 1994 | Toronto, Canada |  | 309 | **67.0** | **62.6** | 64.8 |
| Nanda | 250 | 1995 | Oklahoma, USA |  | 50 | **65.11** | **61.45** | 63.28 |
| Hall | 251 | 1996 | Forest Grove, OR, USA |  | 201 | **62.7** | 60.0 | 61.35 |
| Cole | 252 | 1997 | Tulane, LA, USA |  | 104 | **61.57** | **60.15** | 60.86 |
| Barretto | 253 | 1999 | Michigan, USA |  | 65 | **65.15** | **61.47** | 63.3 |
| Pivnick | 254 | 1999 | Tennessee, USA |  | 391 | **62** | **60** | 61 |
| MacLachlan | 41 | 2002 | Ithaca, NY, USA |  | 13 | **62.21** | **59.99** | 61.11 |
| Bradtmiller | 255 | 2004 | USA |  | 1,886 | **64.00** | **61.17** | 62.6 |
| Dodgson | 4 | 2004 | USA |  | 2,302 | 63.35 | 60.65 | **61.99** |
| Keefe | 256 | 2012 | Canada (Forces) |  | 2,191 | **63** | **59** | 61 |
| Gordon | 257 | 2016 | US Pilots, 977 m, 42 f | 88% Caucasian | 1,372 | **63.4** | **60.3** | 61.85 |
| Parciak | 7 | 2017 | Riverside, CA, USA |  | 120 | **77.4** | **74.8** | 76.1 |
| DOWN SYNDROME |  |  |  |  |  |  |  |  |
|  |  |  |  | M=F-0.1 |  |  |  |  |
| Lyle | 258 | 1972 | Waterloo, Ontario, Canada |  | 22 | **54.3** | **55.6** | 55.0 |
| Jaeger | 246 | 1980 | Philadelphia, PA, USA |  | 50 | **58.6** | **57.4** | 58.0 |
| Krinsky-McHale | 259 | 2014 | New York City, NY, USA |  | 5 | 56.35 | 56.45 | **56.4** |
| AFRICAN AMERICAN |  |  |  | M=F+3.32 |  |  |  |  |
| Herskovits | 260 | 1926 | New York City, USA |  | 529 | **68.1** | 64.85 | 66.5 |
| Davenport | 239 | 1929 | Jamaica |  | 100 | **70.60** | **66.68** | 68.64 |
| Cesario | 5 | 1984 | Washington DC, USA |  | 50 | **62.86** | **59.39** | 61.1 |
| Murphy | 261 | 1990 | Virginia, USA |  | 100 | **66.3** | **62.6** | 64.5 |
| Latta | 6 | 1991 | Tennessee, USA |  | 55 | **66** | **63** | 64.5 |
| Cole | 252 | 1997 | Tulane, Louisiana |  | 236 | **64.63** | **63.2** | 63.92 |
| Barretto | 253 | 1999 | Michigan, USA |  | 61 | **68.97** | **65.93** | 67.5 |
| Pivnick | 254 | 1999 | Tennessee, USA |  | 52 | **70** | **66** | 68 |
| Bradtmiller | 255 | 2004 | USA |  | 1,223 | **67.70** | **65.51** | 66.6 |
| Dodgson | 4 | 2004 | USA |  | 1,376 | 67.2 | 64.0 | **65.62** |
| Parciak | 7 | 2017 | Riverside, CA, USA |  | 120 | **82.7** | **77.6** | 80.15 |
| NATIVE AMERICAN |  |  |  |  |  |  |  |  |
| Dodgson | 4 | 2004 | USA |  | 26 | 66.1 | 64.1 | **65.12** |
| MULTIPLE ETHNICITIES |  |  |  |  |  |  |  |  |
|  |  |  |  | M=F+3.07 |  |  |  |  |
| Gordon | 262 | 2014 | US Army |  | 6,068 | **64.0** | **61.7** | 62.85 |
| Murray | 263 | 2017 | North Carolina, USA |  | 416 | **65.32** | **61.53** | 63.4 |

| LATINO / HISPANIC |  |  |  | M=F+2.01 |  |  |  |  |
| --- | --- | --- | --- | --- | --- | --- | --- | --- |
| Pryor | 36 | 1969 | Oaxaca, Mexico |  | 106 | **65.0** | **64.0** | 64.5 |
| Kawagoe | 264 | 1998 | Sao Paulo, Brazil |  | 30 | 62.3 | 60.3 | **61.3** |
| Bradtmiller | 255 | 2004 | USA |  | 538 | **64.66** | **62.17** | 63.42 |
| Dodgson | 4 | 2004 | USA |  | 125 | 64.5 | 62.5 | **63.54** |
| Gomes | 265 | 2006 | Uberlandio, Brazil |  | 81 | 70.1 | 68.1 | **69.1** |
| Roehe | 266 | 2008 | Recife, Brazil |  | 135 | 65.2 | 63.2 | **64.2** |
| Arenas | 267 | 2014 | Bogota, Colombia |  | 1,262 | 63.58 | 61.58 | **62.58** |
| Da Cunha | 268 | 2015 | Santa Rosa, Brazil |  | 160 | **65.02** | **62.47** | 63.75 |
| Flament | 82 | 2020 | NJ, USA; Mexico City |  | 329 | 66.5 | **64.5** | 65.5 |

**Supp. Table 2.**
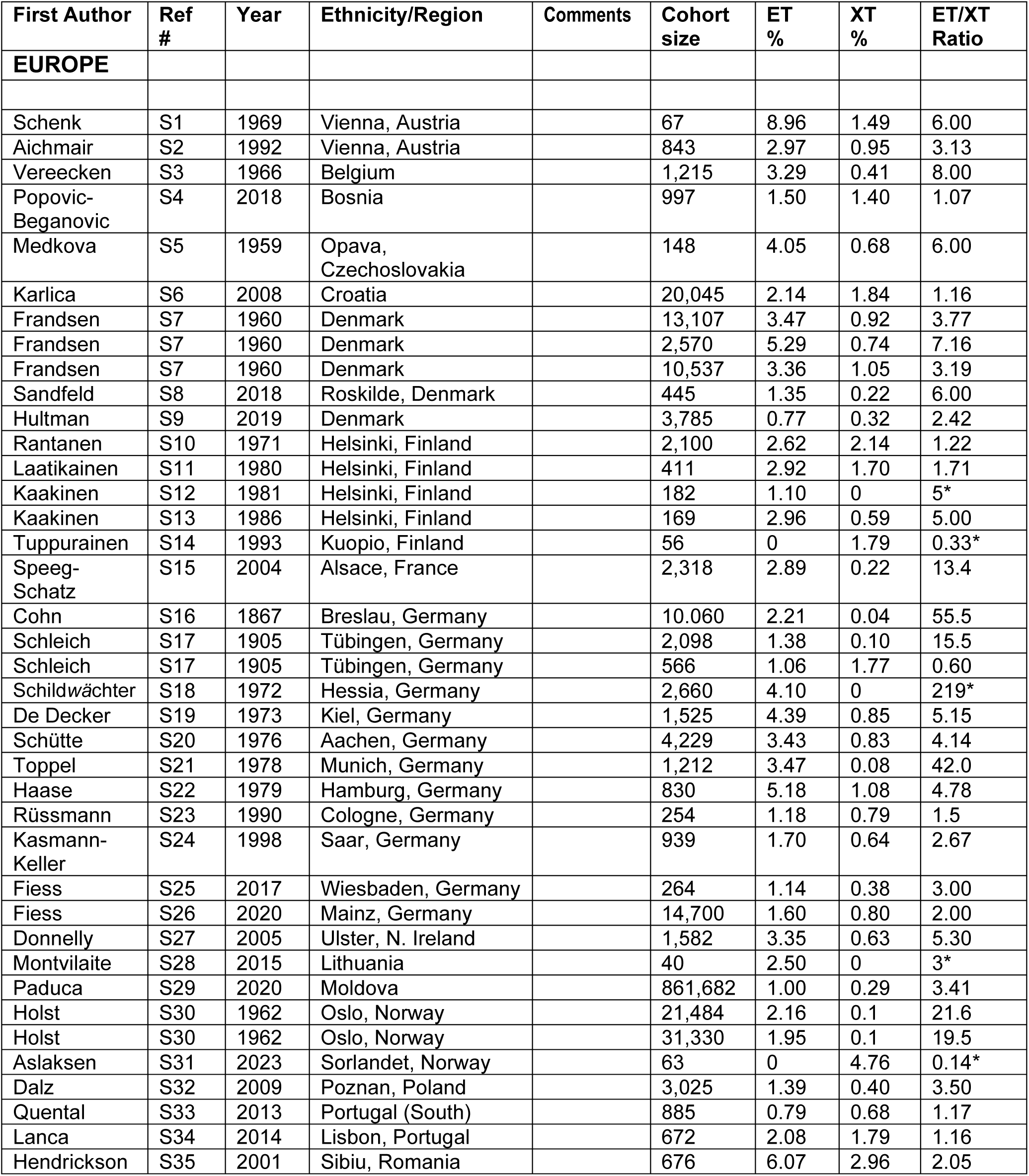
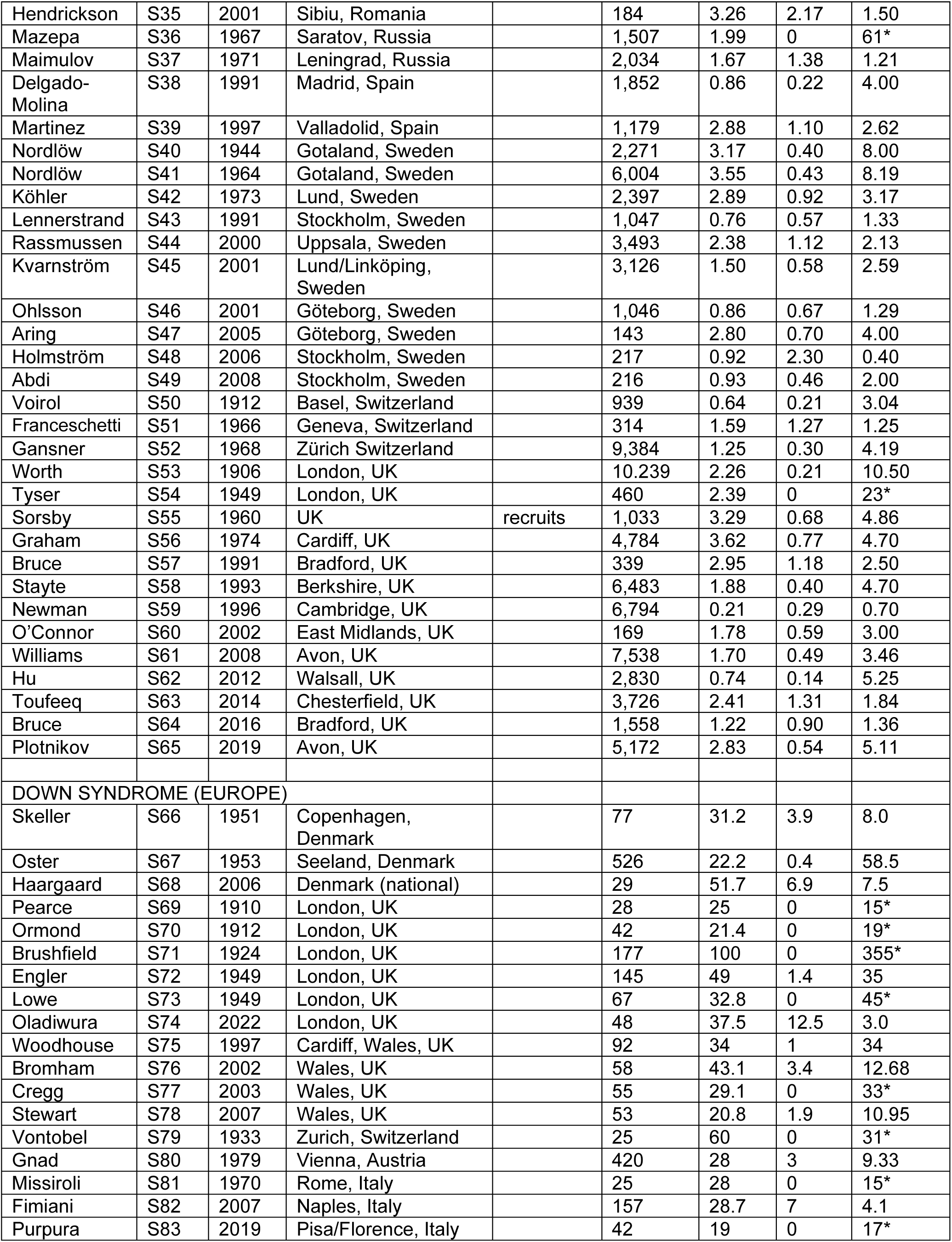
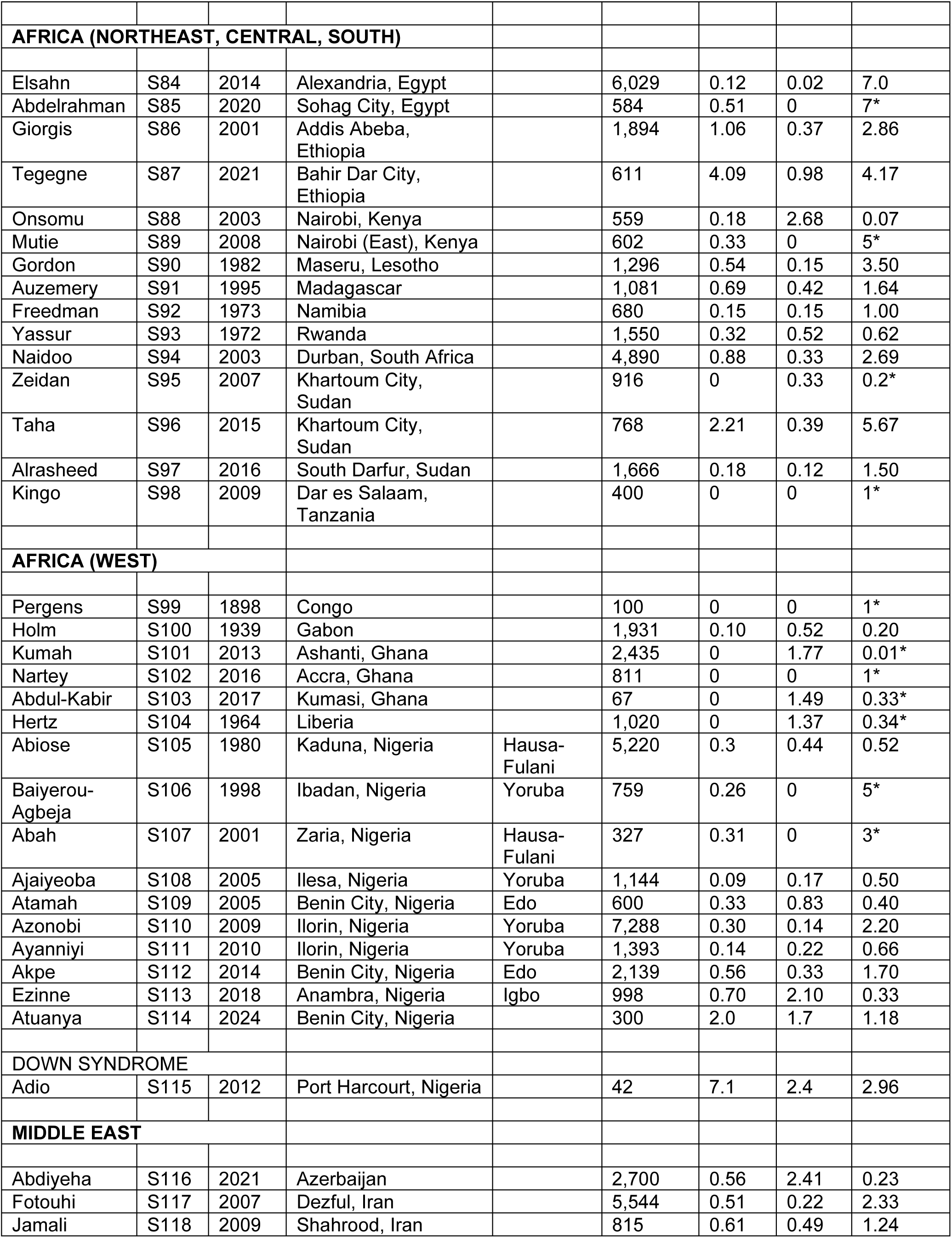
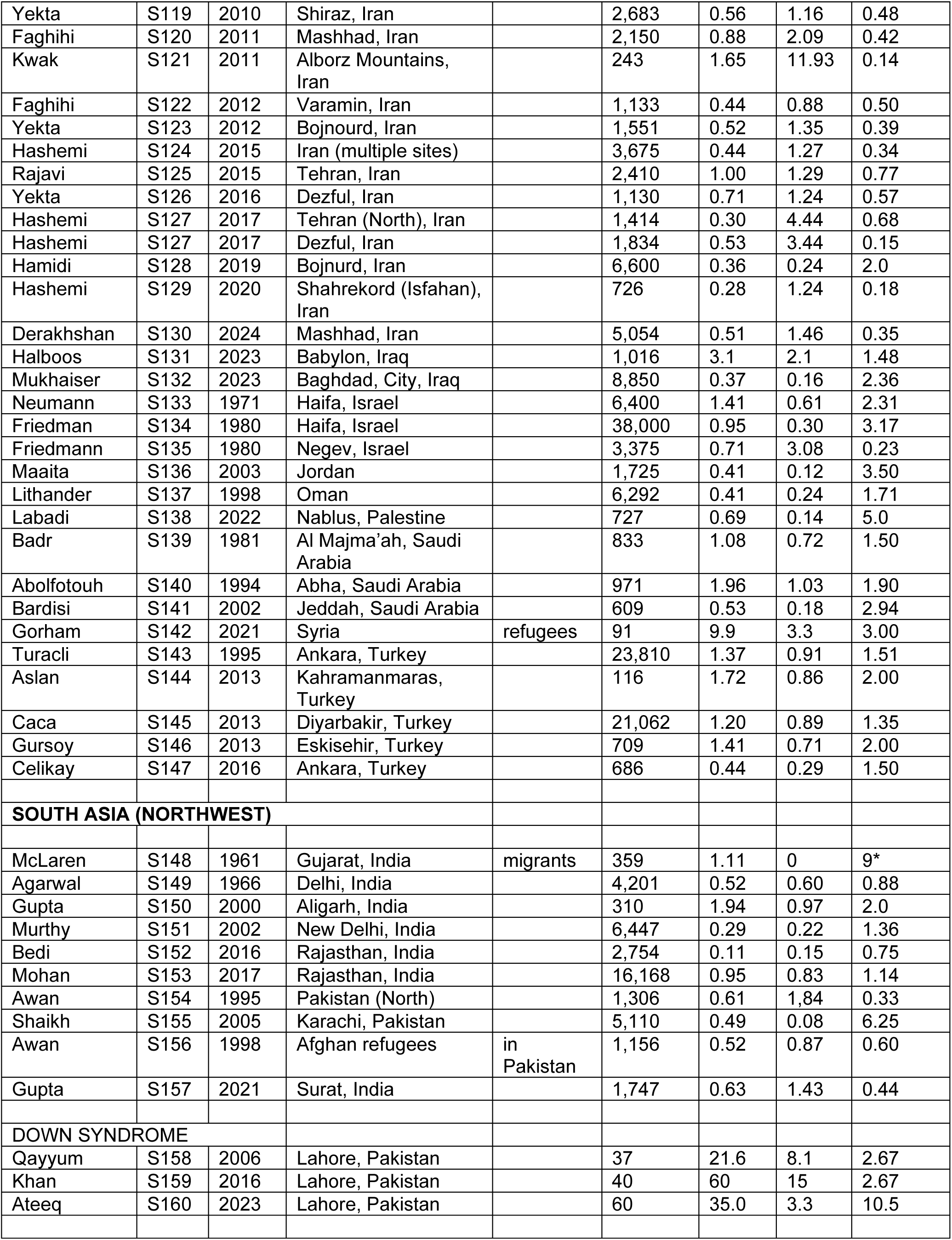
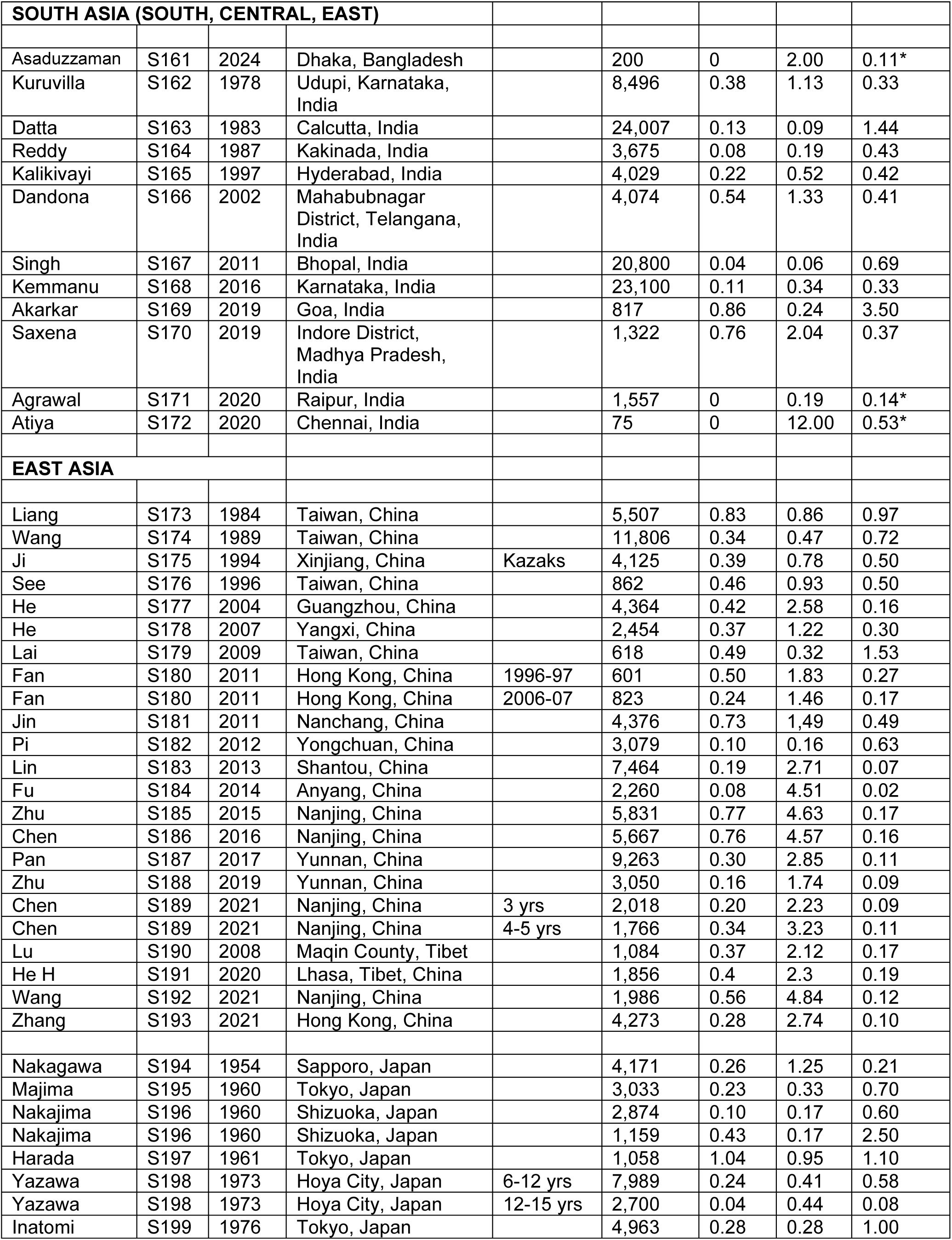
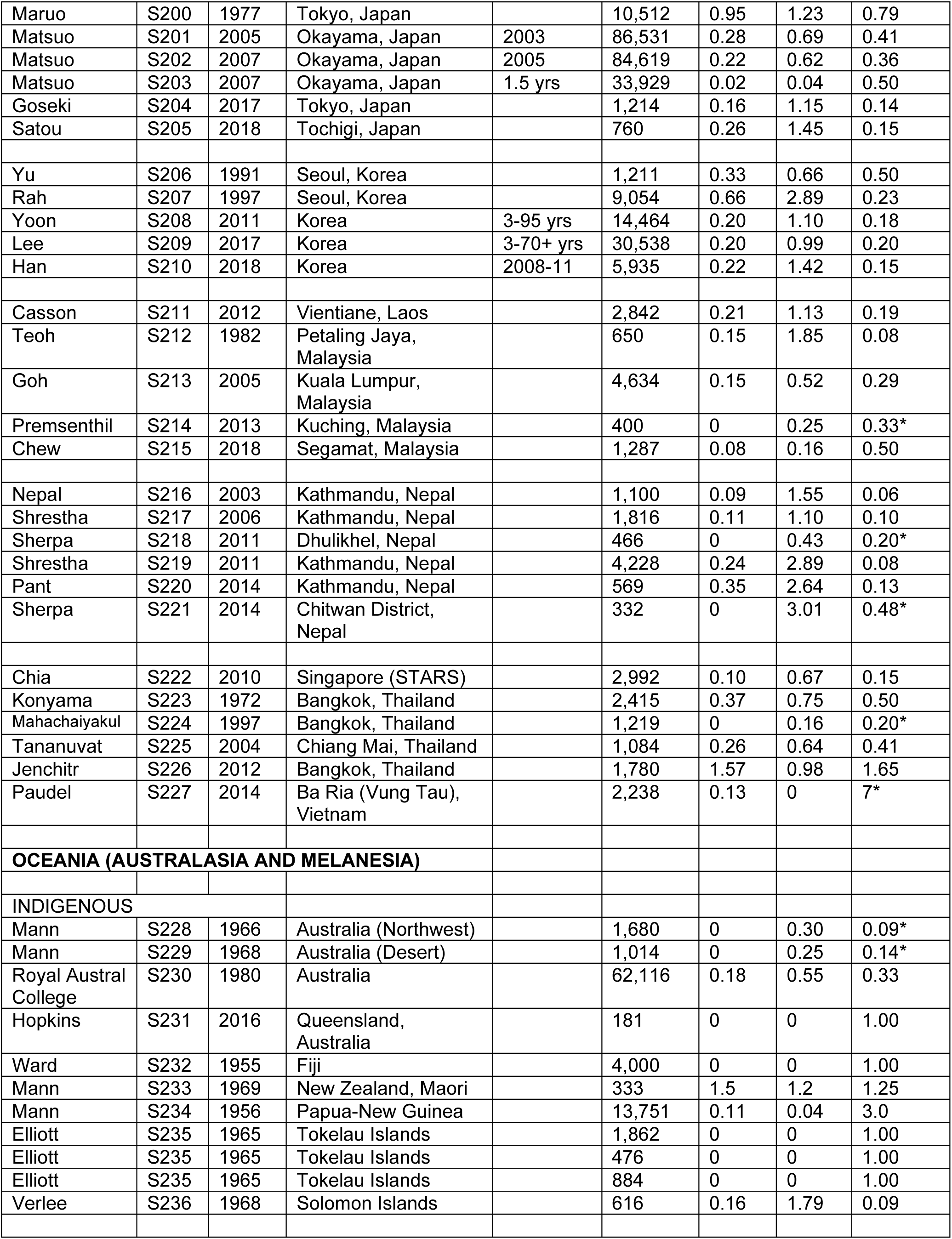
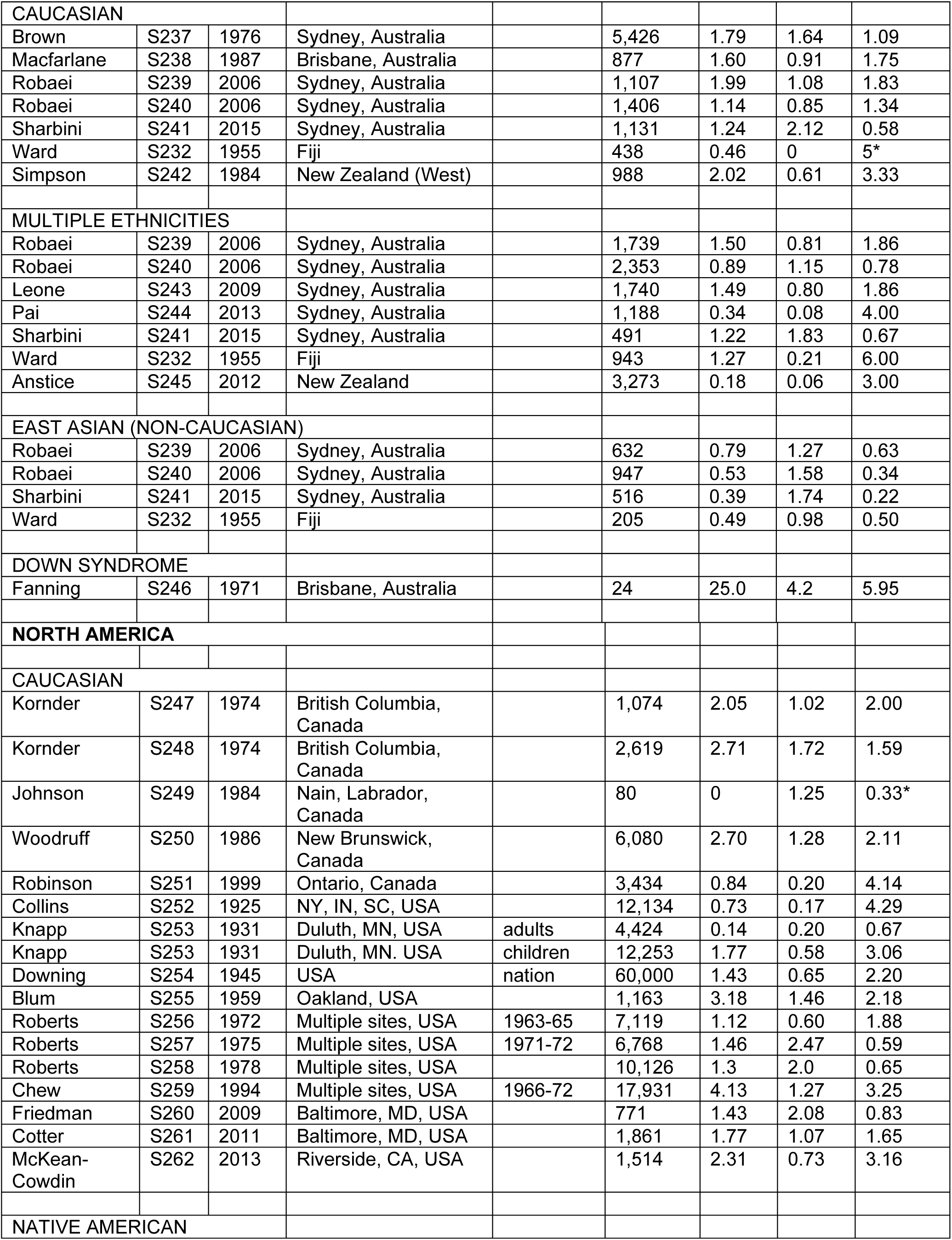
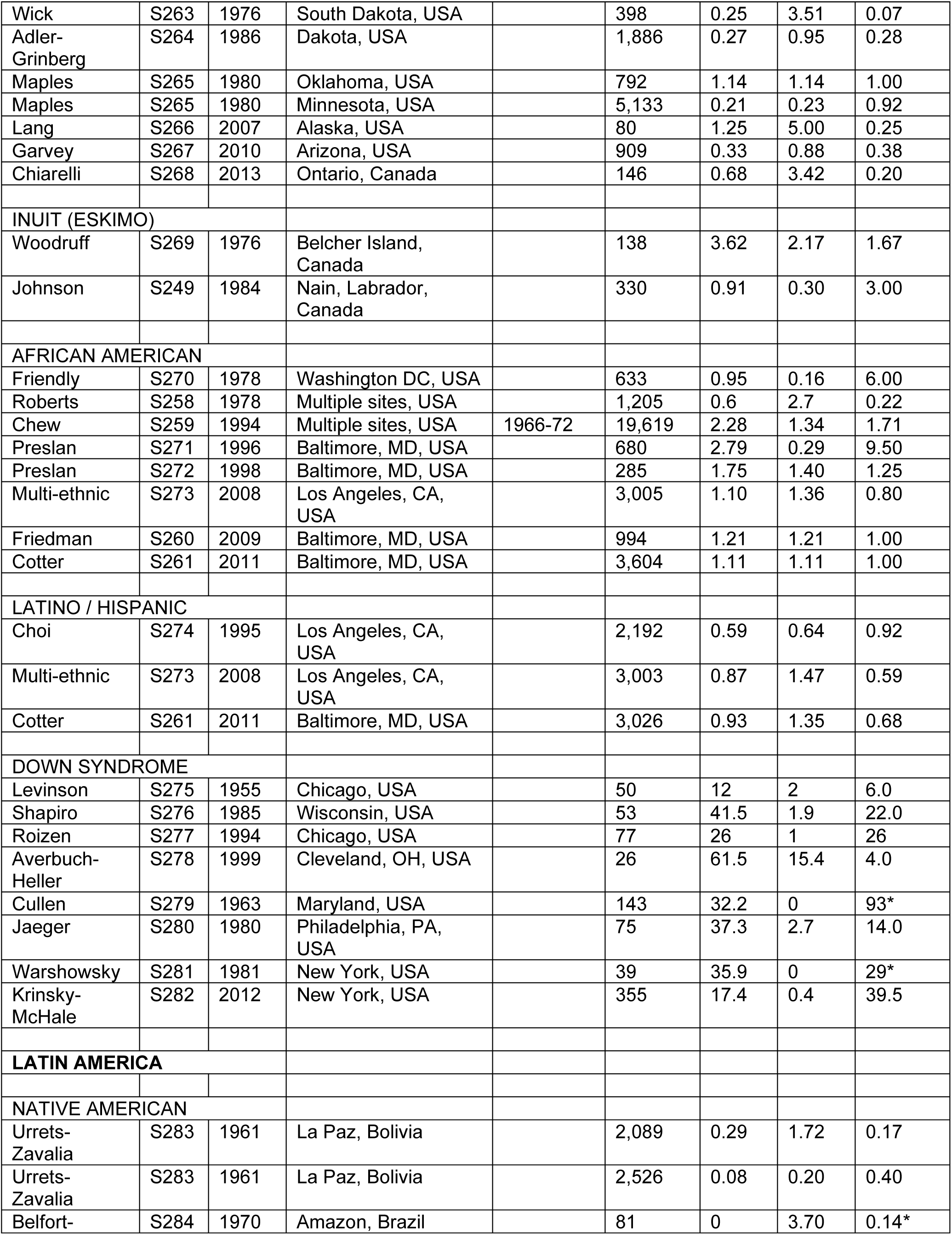
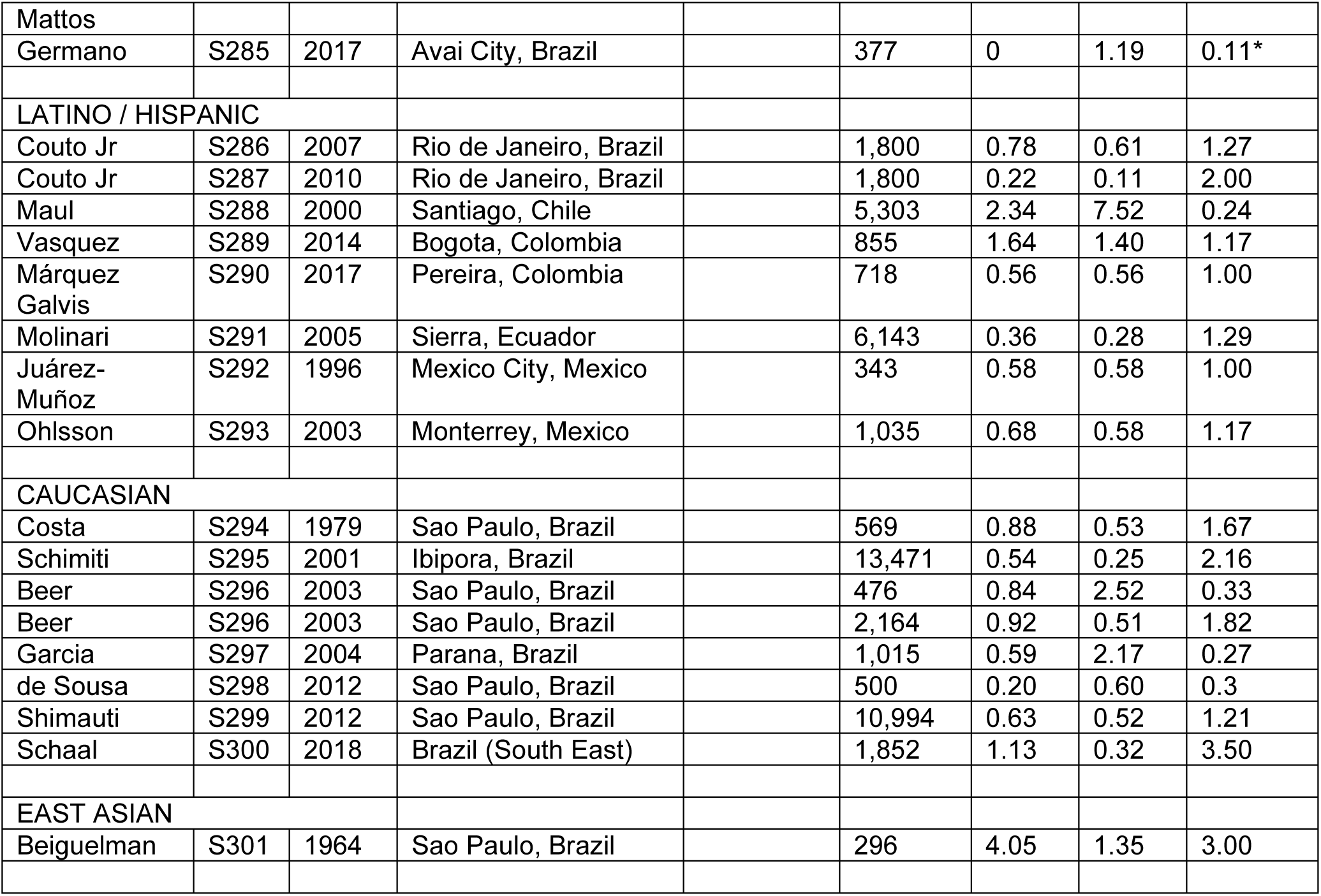
Esotropia/exotropia (ET/XT) ratio in different populations/regions. Data from von Bartheld et al., 2025^22^ (for References – see Supplemental Table S3) * 0.5 was added to each value for the ET/XT ratio when the value was 0, to avoid a zero in the denominator or enumerator, according to Sweeting et al., 2004. Ref, reference number from the Supplemental Table 3.

**Supplemental Table S3: List of References for the ET/XT ratio (from von Bartheld et al., 2025)^22^**

---

**Supplemental Table S4:**
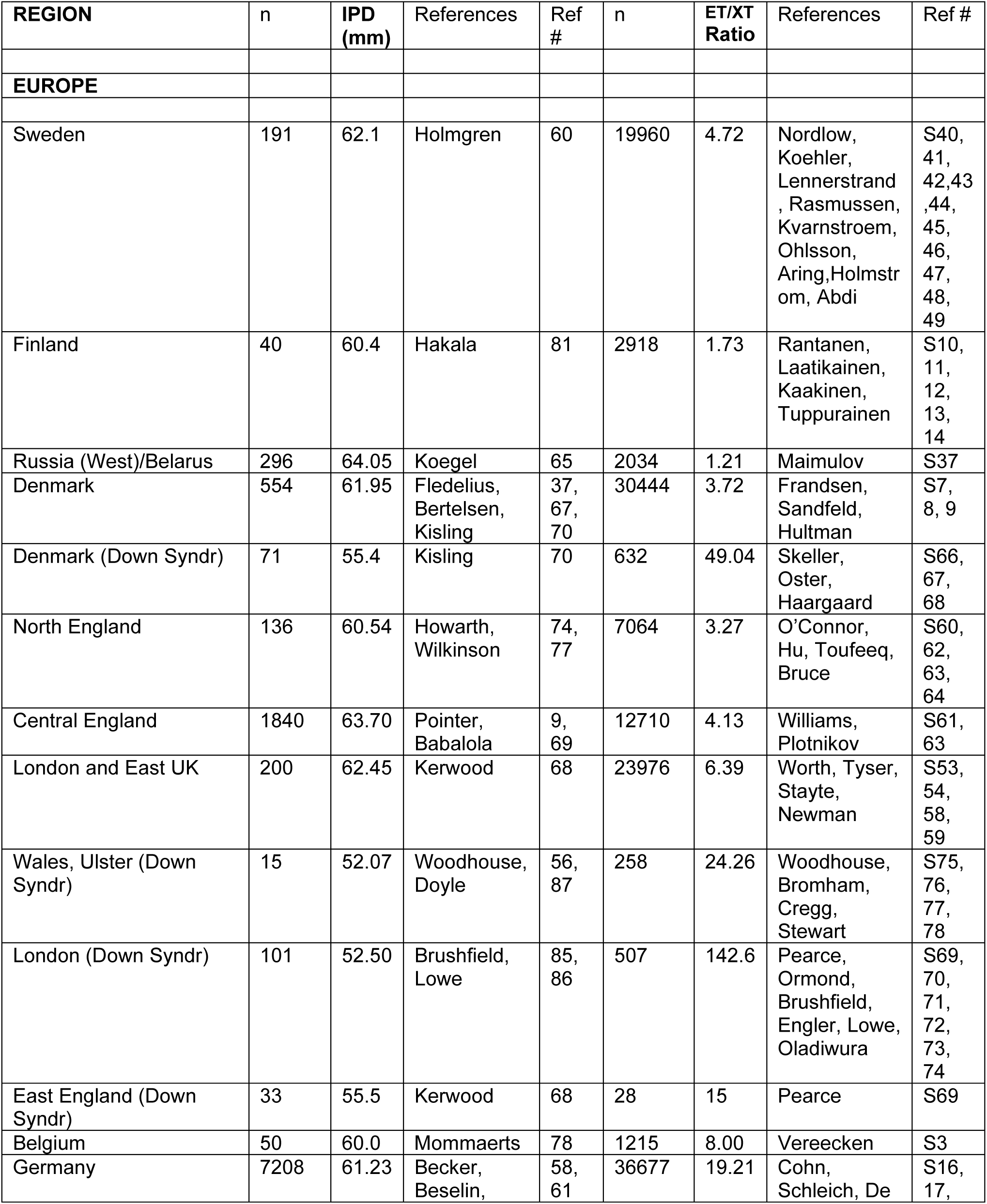
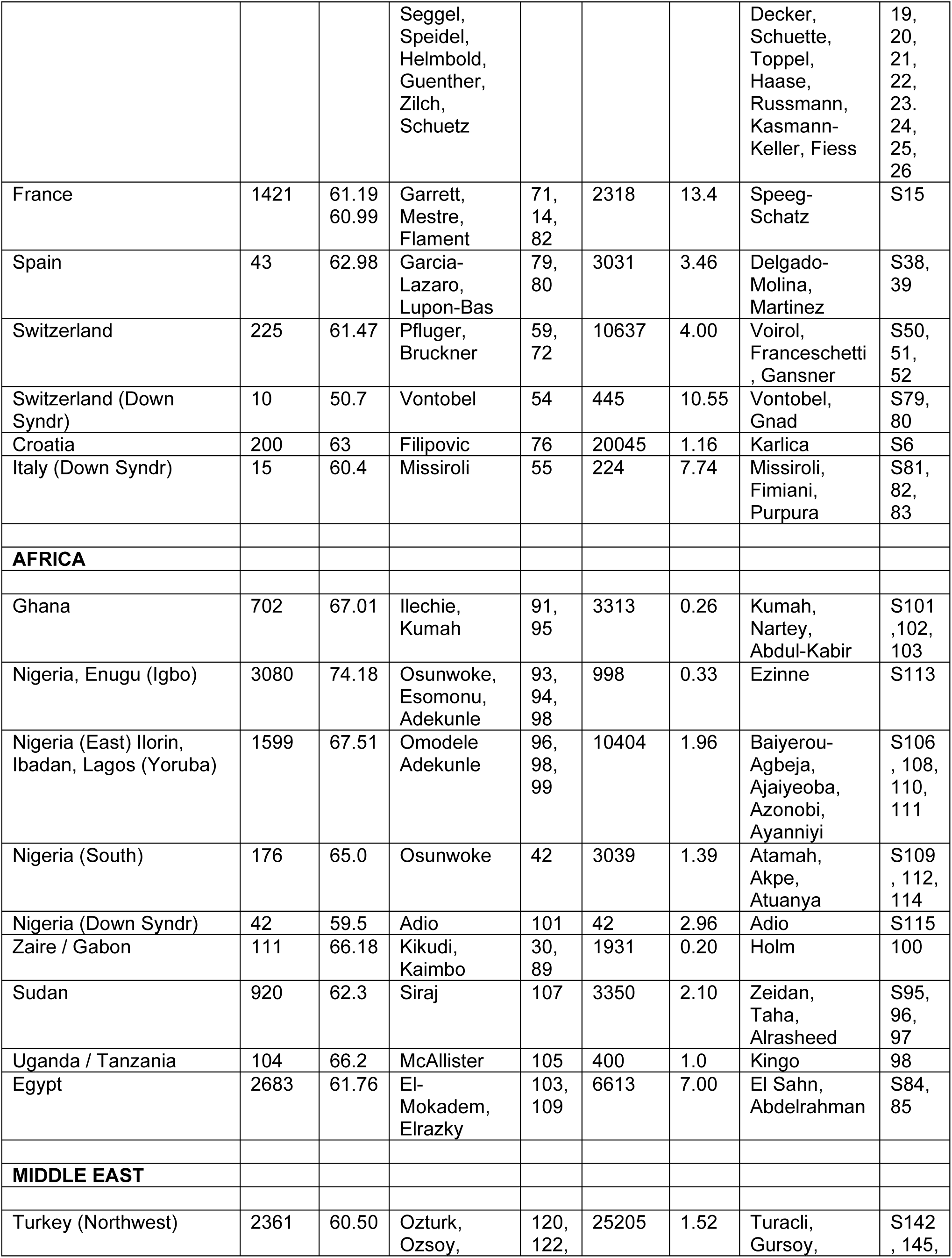
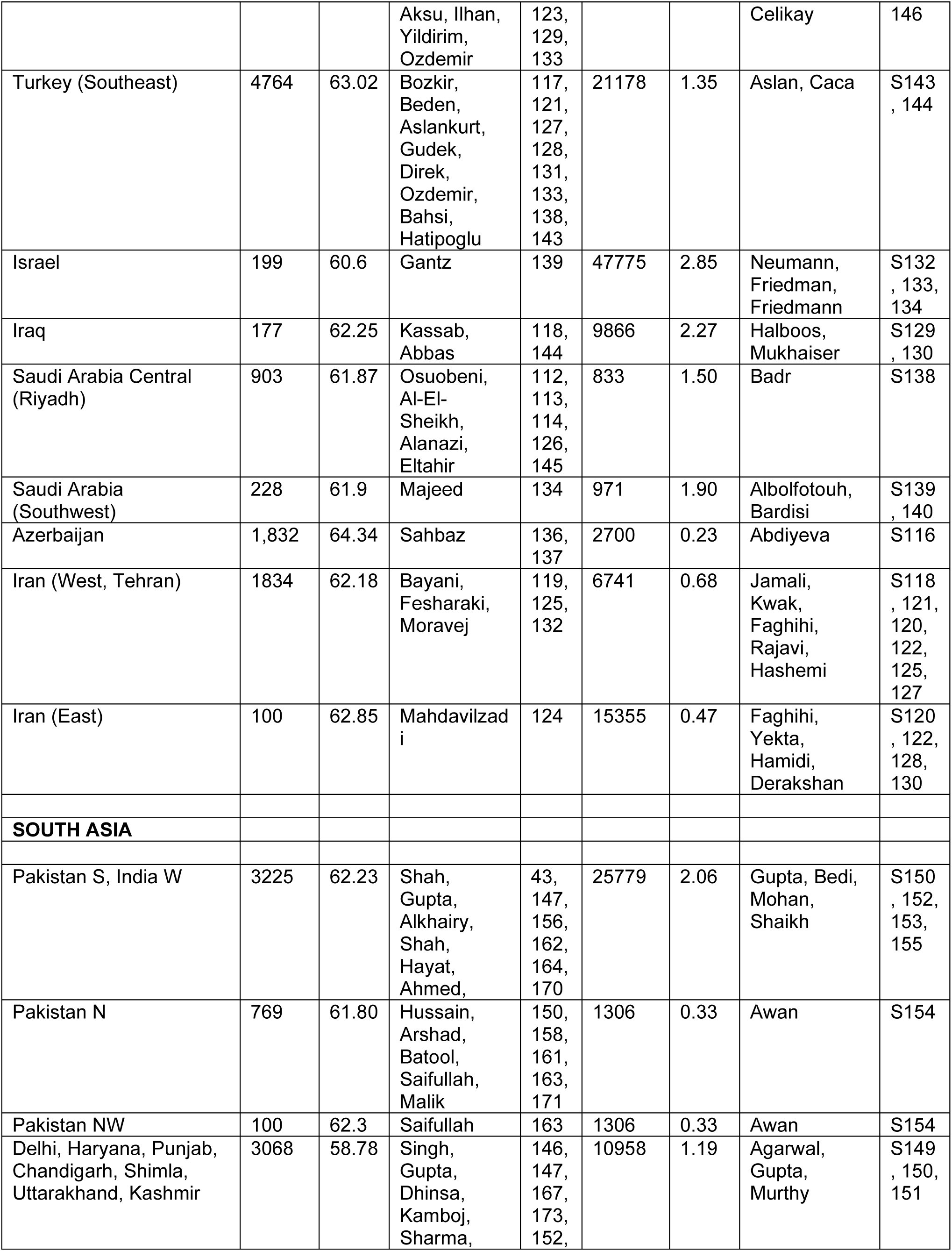
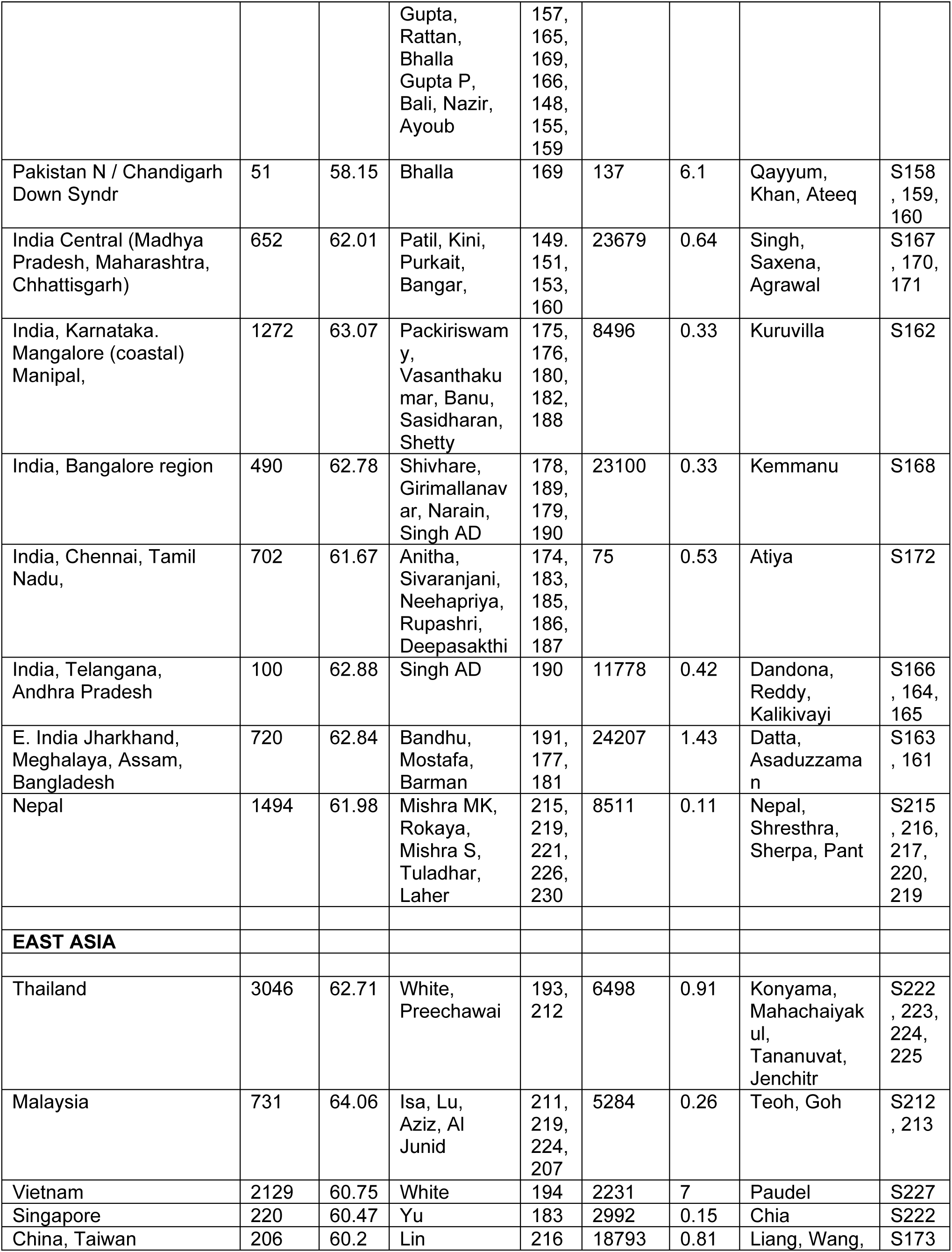
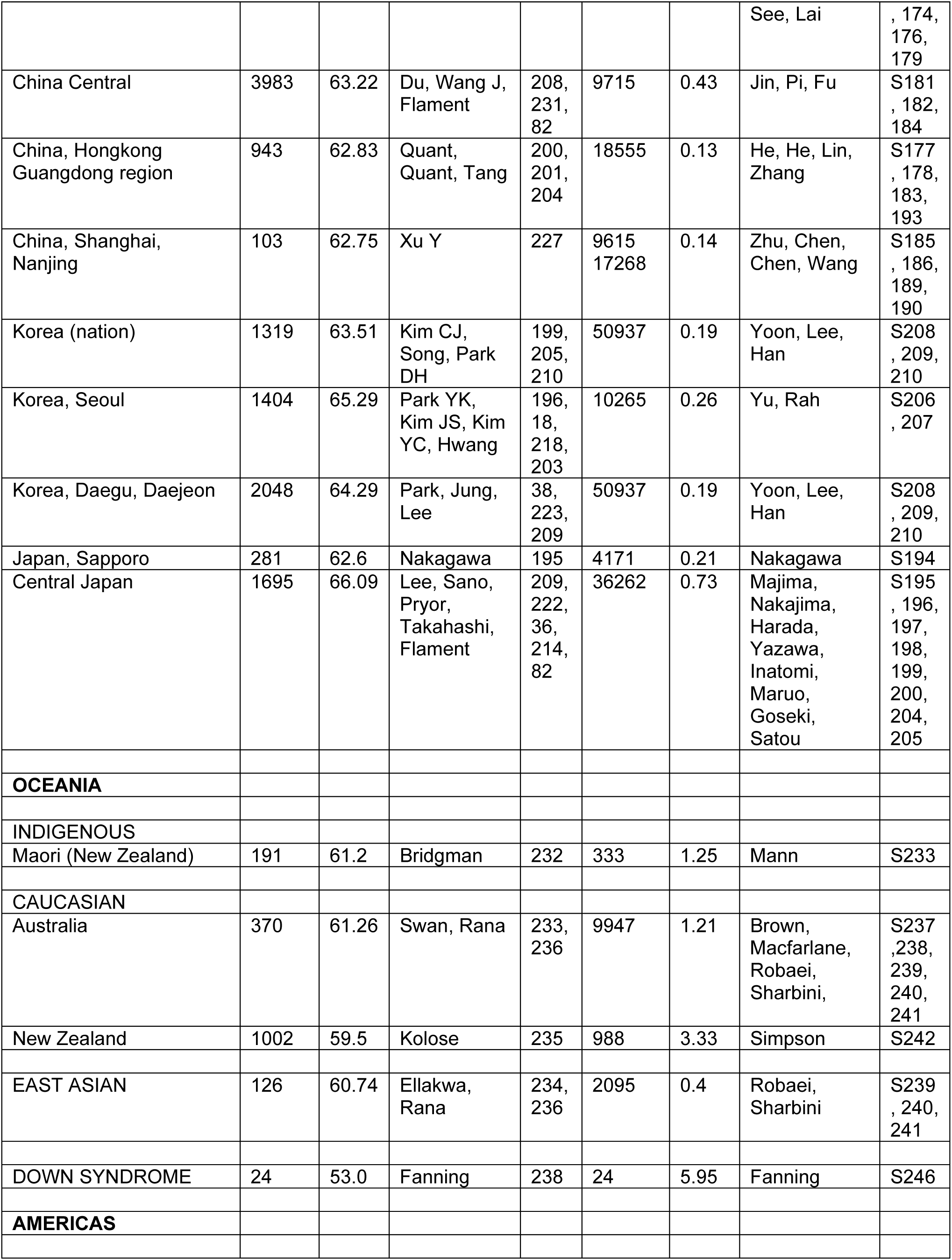
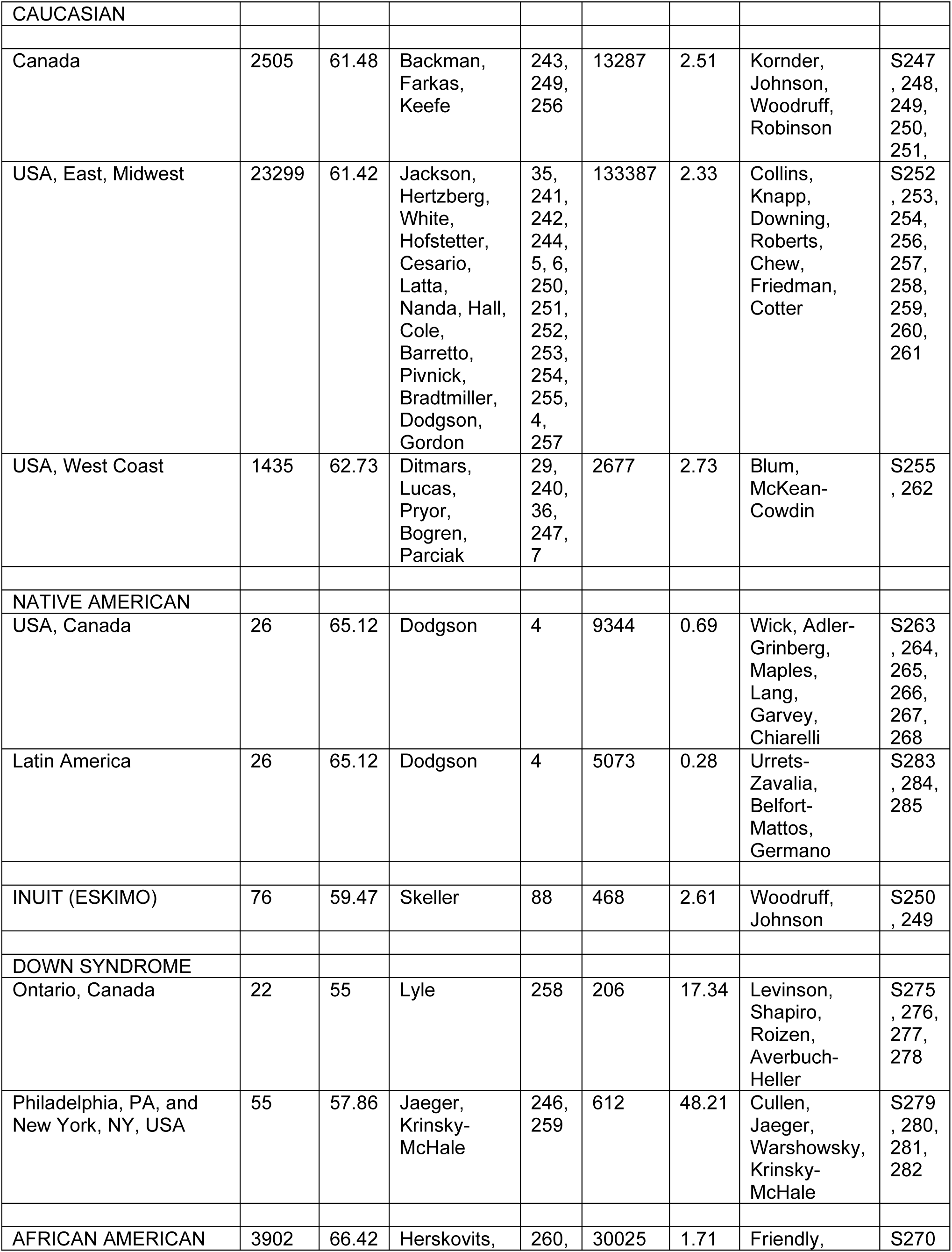
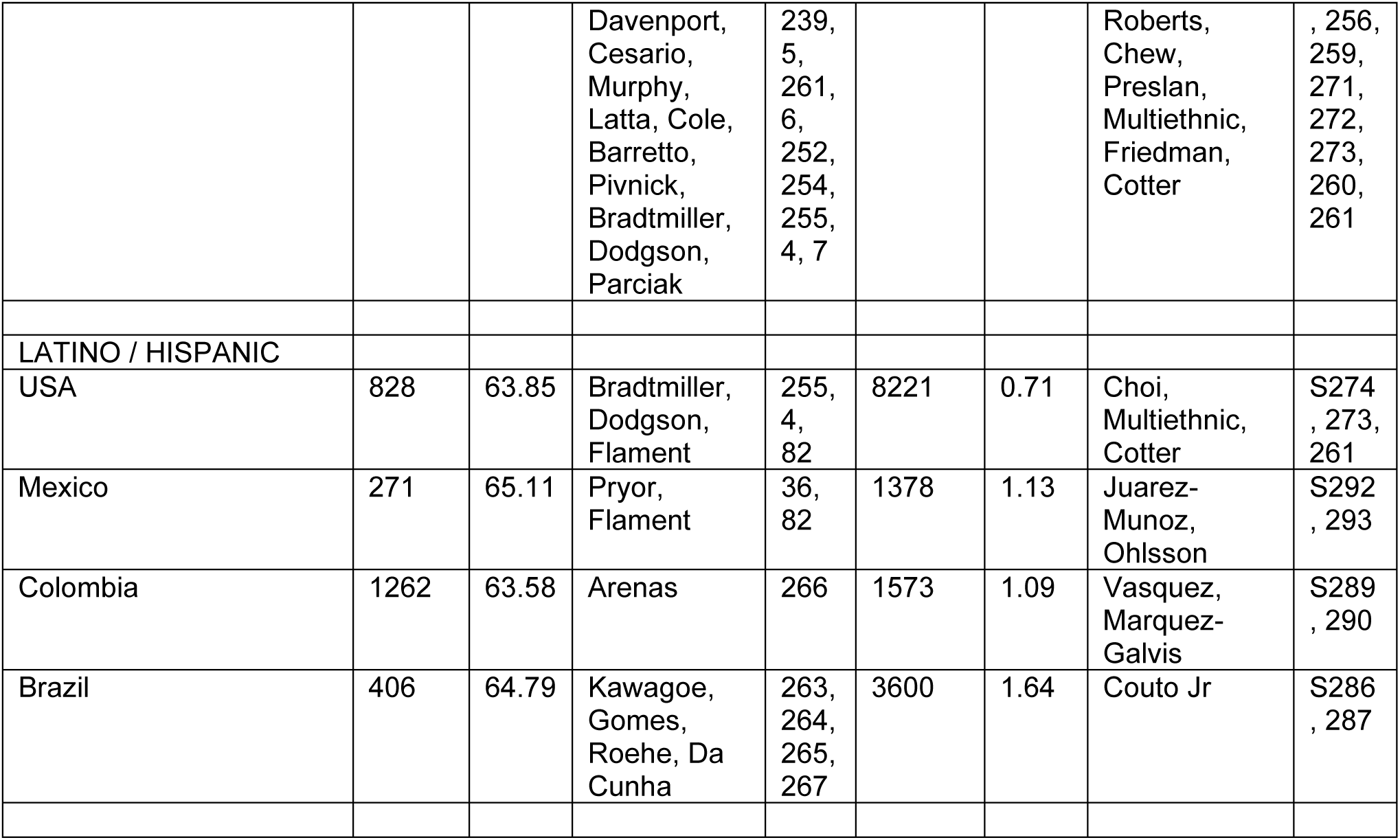
Pairs of ET/XT ratio and IPD within populations for regression analysis.

**Supplemental Table S5:**
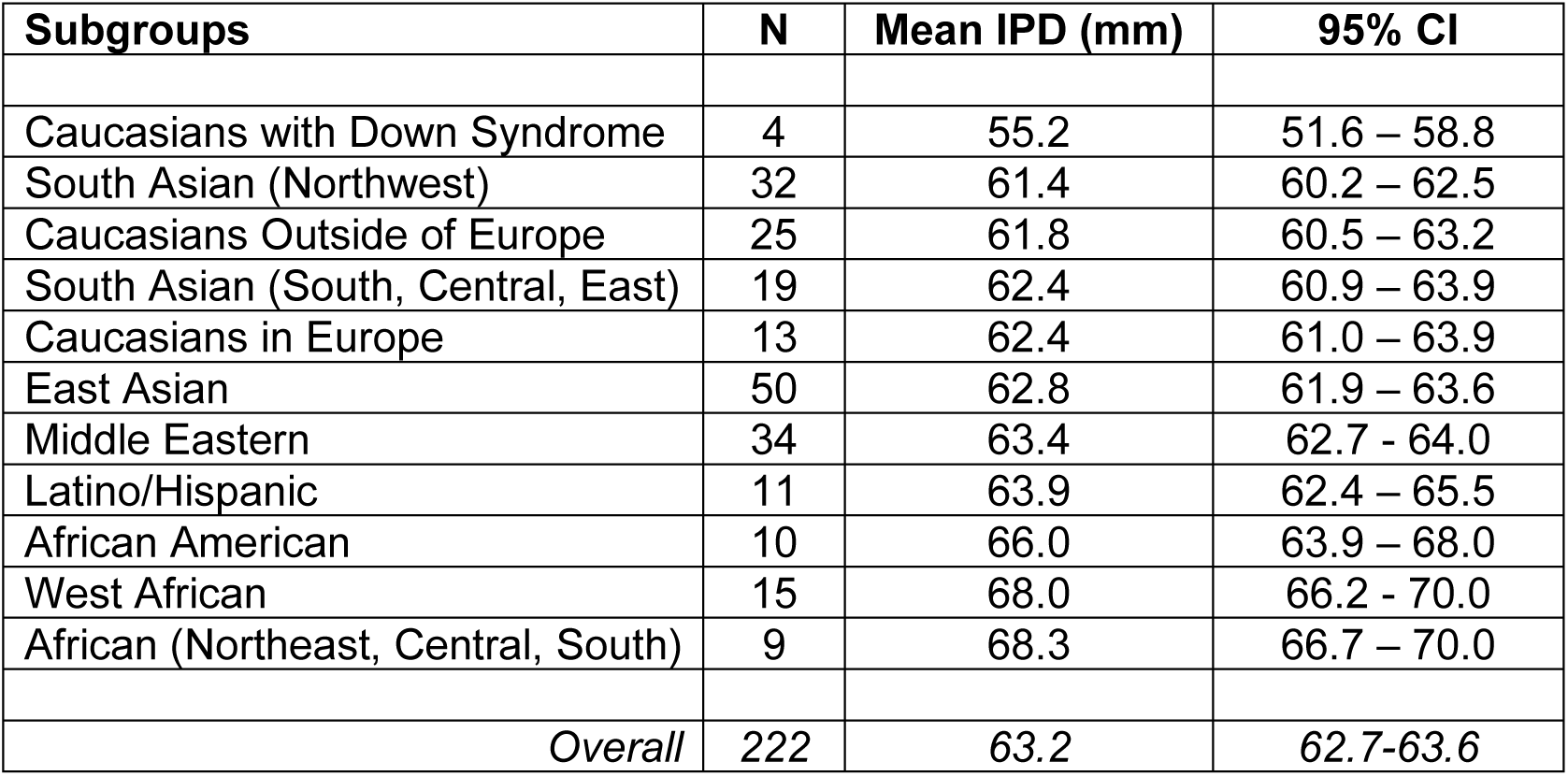
Random pooled estimates of mean IPDs for major ethnicities with 95% confidence intervals (N=222 studies)

